# RNA Sequencing-Based Single Sample Predictors of Molecular Subtype and Risk of Recurrence for Clinical Assessment of Early-Stage Breast Cancer

**DOI:** 10.1101/2021.12.03.21267116

**Authors:** Johan Staaf, Jari Häkkinen, Cecilia Hegardt, Lao H Saal, Siker Kimbung, Ingrid Hedenfalk, Tonje Lien, Therese Sørlie, Bjørn Naume, Hege Russnes, Rachel Marcone, Ayyakkannu Ayyanan, Cathrin Brisken, Rebecka R. Malterling, Bengt Asking, Helena Olofsson, Henrik Lindman, Pär-Ola Bendahl, Anna Ehinger, Christer Larsson, Niklas Loman, Lisa Rydén, Martin Malmberg, Åke Borg, Johan Vallon-Christersson

## Abstract

**Background:** Multigene expression assays for molecular subtypes and biomarkers can aid clinical management of early invasive breast cancer (IBC). Based on RNA-sequencing we aimed to develop robust single-sample predictor (SSP) models for conventional clinical markers as well as molecular intrinsic subtype and risk of recurrence (ROR) that provide clinically relevant prognostic stratification.

**Methods:** A uniformly accrued breast cancer cohort of 7743 patients with RNA-sequencing data from fresh tissue was divided into a training set (n=5250) and a reserved test set (n=2412). We trained SSPs for PAM50 molecular subtypes and ROR assigned by nearest-centroid (NC) methods and SSPs for conventional clinical markers from histopathology data. Additionally, SSP classifications were compared with Prosigna in two external cohorts (ABiM, n=100 and OSLO2-EMIT0, n=103). Prognostic value was assessed using distant recurrence-free interval (DRFi).

**Results:** In the test set, agreement between SSP and NC classifications for PAM50 (five subtypes) and Subtype (four subtypes) was high (85%, Kappa=0.78) and very high (90%, Kappa=0.84) respectively. Accuracy for ROR risk category was high (84%, Kappa=0.75, weighted Kappa=0.90). The prognostic value for SSP and NC classification was assessed as equivalent and added clinically relevant prognostic information. Agreement for SSP and histopathology was very high or high for receptor status, while moderate and poor for Ki67 status and Nottingham histological grade, respectively. SSP concordance with Prosigna was high for subtype (OSLO 83% and ABiM 80%, Kappa=0.73 and 0.72, respectively) and moderate and high for ROR risk category (68% and 84%, Kappa=0.50 and 0.70, weighted Kappa=0.70 and 0.78). In pooled analysis, concordance between SSP and Prosigna for emulated treatment recommendation dichotomized for chemotherapy (yes vs. no) was high (85%, Kappa=0.66). In postmenopausal ER+/HER2-/N0 patients SSP application suggested changed treatment recommendations for up to 17% of patients, with nearly balanced escalation and de-escalation of chemotherapy.

**Conclusions:** Robust SSP models, mimicking histopathological variables, PAM50, and ROR classifications can be derived from RNA-sequencing that closely matches clinical tests. Agreement and DRFi analyses suggest that NC and SSP models are interchangeable on a group-level and nearly so on a patient level. Retrospective evaluation in ER+/HER2-/N0 IBC suggested that molecular testing could lead to a changed therapy recommendation for almost one-fifth of patients.

## Introduction

The majority of women with early-stage invasive breast cancer (IBC) are candidates for adjuvant systemic treatment. Prognosis and treatment decisions are routinely based on menopausal status, disease burden, Nottingham histological grade (NHG), and immunohistochemical (IHC) measurements of estrogen receptor (ER), progesterone receptor (PR), human epidermal growth factor receptor 2 (HER2), and the proliferation marker protein Ki67, as well as the copy number of HER2 assessed by *in situ* hybridization ^1^. Diverse prognosis and unpredictable benefits of adjuvant treatment are prominent in the large ER+/HER2-luminal subgroups of breast cancer (BC). Here, overtreatment remains a major clinical challenge, a cause of decreased quality of life, and a high economic burden for the individual and society.

Multigene expression assays have in the past decades been demonstrated to provide guidance in the selection of patients with luminal disease for adjuvant chemotherapy in addition to endocrine treatment, especially in postmenopausal patients ^2-4^. Whereas most multigene signatures are developed by the public research community, clinical use has largely been restricted to commercial implementations of individual signatures using targeted assays ^5^. These clinical tests are based on data from mainly retrospective analyses of different patient cohorts, but also on a few prospective clinical trials ^2,6^. An important limitation of current clinical multigene tests is their targeted design, providing only a limited number (typically one) of clinically useful outputs per analysis. In this context, global mRNA sequencing (RNA-sequencing) may provide a more generic solution, but current prediction models lack validation.

One of the current targeted clinical multigene tests is the Prosigna assay, which is based on the PAM50 molecular subtype classification ^7^, omitting the Normal-like subtype, and implemented on the Nanostring nCounter Analysis System. Along with the PAM50 subtypes, Parker and colleagues also reported the construction of risk of recurrence (ROR) scores based on subtype correlations and a dichotomized tumor size variable ^7^. The equation for ROR and risk classification cutoff was constructed using a cohort of predominantly node-negative patients not receiving adjuvant systemic therapy and with long (median 9 years) clinical follow-up, while prediction of preoperative chemotherapy sensitivity was evaluated in patients based on pathological complete response ^7^. A re-engineered assay based on PAM50 classification and developed for formalin-fixed, paraffin-embedded (FFPE) tissue was subsequently implemented on the Nanostring nCounter Analysis System and validated as the clinical Prosigna test that reports four subtypes and a ROR score ^8^. Since the first report of the PAM50 subtypes and ROR, the prognostic value of these classifications has repeatedly been demonstrated ^9-12^ and it has been shown that the Prosigna test recapitulates and matches properties of the published PAM50 classifier and ROR model ^8,13^.

Similar to most multigene expression models, PAM50 subtypes and ROR rely on normalization to quantify gene expression relative to a reference. New samples are assigned a class label by measuring a distance in relative gene expression space to class centroids and selecting the nearest one, i.e., nearest-centroid (NC) classification. In order for the distance measure to be valid, new samples must be normalized to appropriately adjust their gene expression in relation to the used reference centroids. Failure to do so can result in erroneous classification ^8,14-17^ but when performed correctly classifications are valid ^8^. One strategy is to use a standardized normalization of every new sample to be classified. However, this requires the use of a uniform platform consistent over time, which might be challenging, and methods reliant on data transformations derived from other samples are not considered true single-sample predictors.

An alternative strategy involves models built on rules that only consider gene expression values from a single sample, independent of normalization to reference samples and was suggested for absolute assignment of breast cancer intrinsic molecular subtype (AIMS) by Paquet and Hallett ^14^. Such models are built by identifying a small set, e.g., <50, of gene-pair rules specific for the respective class and based on the form: expression of gene A > expression of gene B. New samples are classified by evaluating these gene-pair rules and assigning a class by the largest number of fulfilled rules or by a probability model ^14,18^. Such models can rightfully be termed single-sample predictors (SSPs) and have been shown to be applicable for cancer classification problems including distinct molecular subtypes ^14,19^ as well as for continuous variables such as cell proliferation signal ^19^. Even though SSP models have features attractive for clinical implementation, robust implementations relevant for BC diagnostics and treatment decision support are still lacking.

In the present study we aimed to develop and benchmark RNA-sequencing based SSP models for conventional clinical BC biomarkers, the four intrinsic molecular subtypes corresponding to Prosigna subtypes, and ROR scores. To construct and evaluate SSPs we used a uniformly accrued population-based cohort of BC comprising 7868 patients from South Sweden analyzed by whole transcriptome RNA-sequencing (>19000 genes) through the Swedish Cancerome Analysis Network - Breast (SCAN-B, ClinicalTrials.gov ID NCT02306096) study ^20-22^. This unique cohort allows generalization and real-world side-by-side prognostic assessment of developed predictors in clinically relevant subgroups with available follow-up data. Moreover, retrospective analysis enables estimation of the possible impact on therapy recommendations from SSP-based molecular subtype and ROR in the clinical decision-making. Finally, we performed benchmarking of our developed SSPs against the Prosigna test in two smaller independent clinical series. Taken together, we demonstrate that appropriate sampling of fresh BC tissue, i.e., not FFPE, can be effectively integrated into current clinical routine practices and used for cost effective RNA-sequencing with different SSPs for expression-based diagnostic and prognostic purposes. Thus, our study moves the usefulness and role of RNA-sequencing one step closer towards clinical implementation in BC, and provides a resource for continued exploration of expression-based BC markers.

## Material and Methods

### Ethics approval and informed consent

All SCAN-B and ABiM enrolled patients provided written informed consent prior to study inclusion. The included ABiM cohort is from patients enrolled in the population-based All Breast Cancer in Malmö study and data is available on line as described ^23^. Ethical approval was given for the SCAN-B study (approval numbers 2009/658, 2010/383, 2012/58, 2013/459 and 2015/277) and ethical approval was given for the ABiM study (approval number 2007/155) by the Regional Ethical Review Board in Lund, Sweden, governed by the Swedish Ethical Review Authority, Box 2110, 750 02 Uppsala, Sweden.

Included normal breast tissue was obtained from women undergoing mammoplasty surgery with no previous history of BC, who gave informed consent, and the tissue samples were examined by the pathologist to be free of malignancy and processed as described ^24^. The study was approved by the Cantonal ethics committee, Commission Cantonale d’éthique de la recherche sur l’être humain, CER-VD, Avenue de Chailly, 1012 Lausanne, Switzerland (Approval number 183/10).

The included OSLO2-EMIT0 breast cancer cohort is from OSLO2, a prospective observational study that enrolled BC patients with primary operable disease at hospitals in southeastern Norway between 2015 and 2020. Informed consent was obtained from all patients included in the OSLO2 study. Ethical approval was given for the OSLO2 study (approval number 29668) by the Norwegian South-East Regional Committee for Medical and Health Research Ethics, Postboks 1130, Blindern, 0318 Oslo, Norway.

### Patient material

Included patient cohorts are outlined in Figure 1a. The SCAN-B material comprises a population-based consecutively enrolled series of BC patients accrued at seven hospitals in the south Sweden health care region, and at two additional Swedish hospitals (Jönköping and Uppsala). Patient management, including adjuvant systemic and radiotherapy treatment have been performed according to national and regional treatment guidelines at the time of enrollment. SCAN-B patients included in this study were enrolled between September 1 2010 and May 31 2018 and sample collection and work-up followed reported SCAN-B procedures and protocols ^20,22^. Clinicopathological and follow-up data as well as information on adjuvant medical treatment was obtained from the Swedish National Quality Register for Breast Cancer (NKBC ^25^). Clinicopathological data reported to NKBC was determined by each respective local pathology department and according to current Swedish clinical guidelines and definitions. For details on pathological assessment see KVAST documents published by the Swedish society of Pathologists (Svensk förening för Patologi – KVAST – document) ^26^. For the earlier part of the material (2010-2014), characteristics of the enrolled patients, collected samples, and RNA-sequencing data has previously been shown to represent the BC population in the recruitment region ^20-22,27^. Available data on adjuvant therapy for the SCAN-B cohort include dichotomized status for systemic endocrine, chemotherapy and HER2-directed therapy. The indication for adjuvant therapy in patients with ER+/HER2-tumors is regularly updated and documented in the Swedish national treatment guidelines.

**Figure 1.**
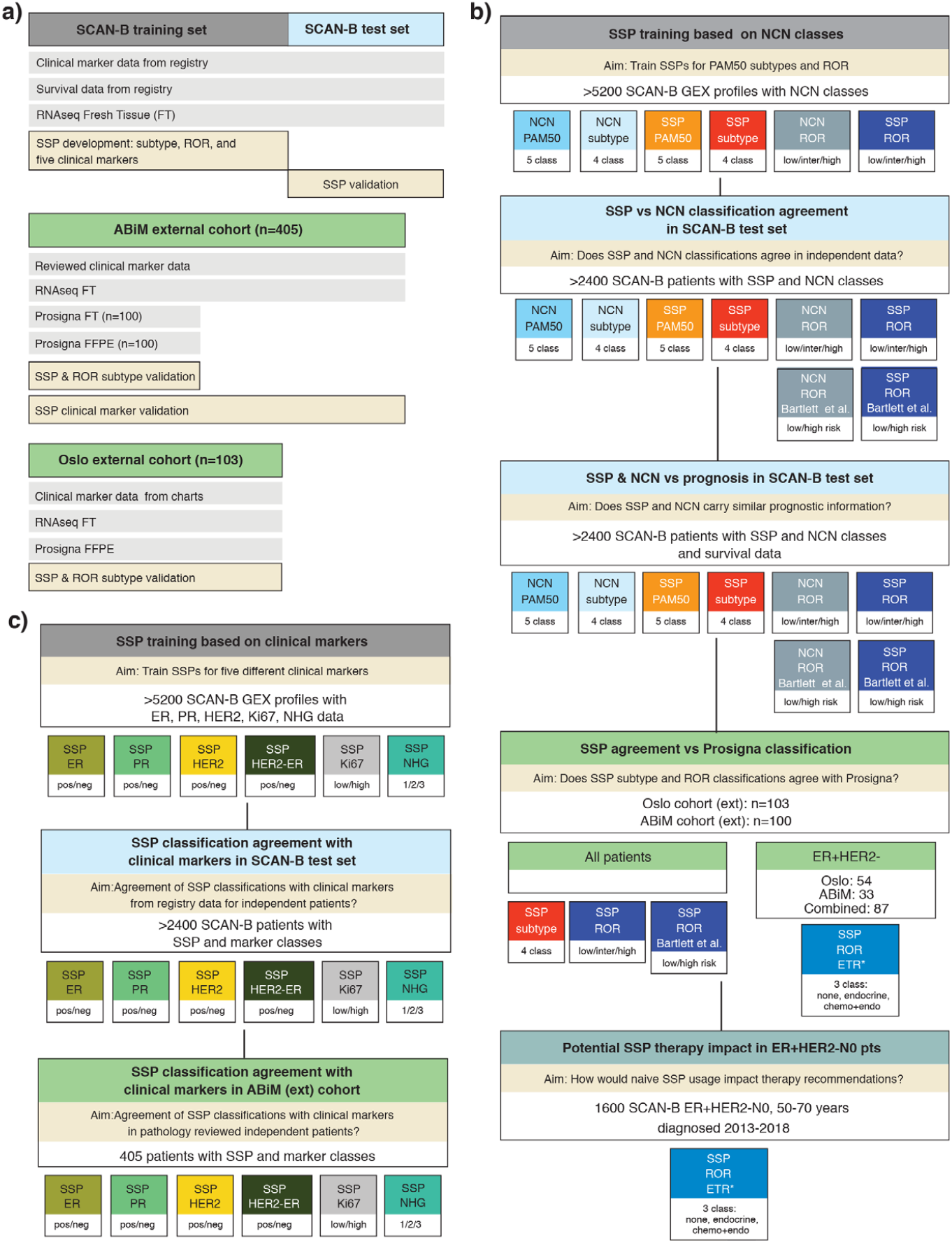
Outline of the study. **(a)** Study cohorts: SCAN-B, ABiM, and OSLO2-EMIT0. Available data types and usage outlined. FT: fresh frozen tissue. **(b)** Scheme for development and validation of SSP models for PAM50 subtypes and ROR based on training versus NCN equivalents. Scheme outlines created SSP models and their usage in different cohorts. NCN and SSP 4-class subtype models include Basal-like, HER2-enriched, Luminal A and Luminal B subtypes. Binary SSP-ROR and NCN-ROR risk classes were created similar as described by Bartlett et al. ^5^. An emulated 3-group SSP-ROR treatment recommendation (SSP-ROR-ETR) was created based on published Norwegian guidelines for Prosigna usage and applied to relevant SCAN-B patients based on guidelines. **(c)** Scheme for development and validation of SSPs for clinical markers: ER, PR, HER2, Ki67, and NHG. Scheme outlines created SSP models and their usage in different cohorts.

The external OSLO2-EMIT0 cohort is a population-based consecutive clinical series of early BC patients accrued during 2015 and 2016 as part of the OSLO2 study ^28^. The external ABiM cohort is a consecutive clinical series of patients with preoperative diagnosis of IBC scheduled for surgery in Malmö, Sweden, during the years 2007–2009 ^29^. For the OSLO2 and ABiM material, freshly collected, macroscopically evaluated, and snap frozen tumor tissue was obtained by clinical pathologists at pathology departments as described ^29,30^ and total RNA was extracted similar to SCAN-B cases. Consensus scoring for the ABiM material from histopathology reassessment has previously been performed as described by Brueffer et al. ^23^.

### Gene expression analysis

RNA-sequencing was performed as described ^20^ or by Illumina stranded TruSeq mRNA protocol, either implemented on KingFisher or on the Illumina NeoPrep system. Expression data (Fragments Per Kilobase per Million reads, FPKM) from stringtie was derived from RNA-sequencing data using an analysis pipeline to align and estimate gene expression values for sequenced samples. The RNA-sequencing analysis pipeline is based on a collection of open source software tools; picard tools ^31^, trimmomatic ^32^, bowtie2 ^33^, hisat2 ^34,35^, stringtie ^36^ with the GRCh38 human genome primary assembly, dbSNP^37^, and GENCODE ^38^ transcriptome model as detailed in Supplemental methods. Entrez ID from the Gencode27 metadata was used as gene identifiers.

### Assigning PAM50 subtype and ROR score using nearest centroid

PAM50 subtype and ROR score were assigned by NC classification following a general and established strategy previously described ^7^. This strategy requires an appropriate static reference set to use for normalization before calculating correlations to the published PAM50 centroids. In order to correctly transform gene expression we selected a static reference set by matching the clinicopathological metadata of the training population from which the centroids were derived ^7^. Moreover, our large dataset permitted us to advance the NC strategy by selecting multiple reference sets, thereby avoiding relying on a single selection. Therefore, the selection procedure was repeated 100 times to create a series of individual static reference sets, each mimicking the original training population. The collection of reference sets was used to construct an extended NC classifier, i.e., a NC classifier utilizing 100 separate normalizations. Herein we refer to this extended NC classifier as NCN. Using multiple reference sets for normalization makes it possible to account for heterogeneity that prevails even within the restricted boundaries set by the target population. Subtype assignment from NCN was done by majority vote whereas ROR score from NCN was calculated using the average of 100 scores, each calculated as described ^3^ (Supplemental methods).

### Training SSP models using the AIMS procedure

For training SSP models with the described AIMS method ^14^ we used scripts available from the AIMS GitHub repository ^39^. Training was largely performed as described by Paquet et al.^14^ but using gene expression data for >19000 genes, a 5-fold cross-validation repeated five times, and evaluating up to 50 selected gene-pair rules. We used weighted rule selection to adjust for differences in size between subsets of data from different library protocols. Evaluation of parameters used was strictly limited to a subset of the training cohort. To this end, the training cohort was partitioned into provisional training/evaluation sets. However, the final training was performed using the full training cohort that, importantly, had no overlap with our reserved test set. Input gene expression in both training and subsequent validations was untransformed expression values as outputted by stringtie for all protein-coding genes from Gencode27 annotated with Entrez ID. Positive controls for normal breast tissue were omitted from all SSP training. Details of the training are outlined in Supplemental methods.

### Prosigna classification

Prosigna results were obtained from FFPE tumor tissue sections from clinical routine procedures as prescribed for the Prosigna assay (Prosigna insert 2017-07 LBL-C0191-09). For the OSLO2-EMIT0 material, the Prosigna assay was run at the local pathology department using the clinical Prosigna assay on the nCounter instrument in Dx mode as described ^40^. For the ABiM material, the Nanostring gene expression data was generated at the Division of Oncology, Lund University using an appropriate code-set, including the genes for the Prosigna assay, and then sent to Nanostring for readout of Prosigna classification results. In addition, for the ABiM material, paired Nanostring gene expression data and readout of classification results by Prosigna models was also obtained from the RNA extracted from fresh macro-dissected tumor tissue used for RNA-sequencing (Figure 1a).

### Survival analysis

Survival analyses were performed in R version 3.6.1 using the survival package with distant recurrence-free interval (DRFi) as primary endpoint and overall survival (OS), recurrence-free interval (RFi) and breast cancer-free interval (BCFi) as additional endpoints (Supplemental methods and ^41^ and ^42^ for cause of death registry). Survival curves were estimated using the Kaplan-Meier method and compared using the log-rank test. Hazard ratios were calculated through univariable or multivariable Cox regression using the coxph R function. In multivariable analyses, tumor size (mm), patient age at diagnosis (binned in 5 year intervals), lymph node status (N+: positive and N0: negative), and NHG were included as covariates. Median follow-up for DRFi in the full test set of early-stage IBC was 8.1 years (range 0.1-10.9). Median follow-up for DRFi in the subset of postmenopausal ER+/HER2-/N0 IBC for the evaluation of prognosis stratified for ROR and molecular subtype (n=772) was 8.0 years (range 0.1-10.7). Median follow-up time in respective two groups for the additional endpoints were: OS 9.4 and 9.7 years, RFi 8.1 and 8.0 years, BCFi 5.4 and 5.6 years. Median and range for follow-up were calculated for patients with no reported events.

### Data availability statement

Gene expression data will be available through Gene Expression Omnibus (GEO) upon acceptance. Clinicopathological data and a complete list of classifications for samples will be available as Supplementary Data Table 1.

**Table 1.** Study material characteristics and clinicopathological variables for SCAN-B.

|  | Early stage follow-up cohort<sup>a</sup> |  | Test set<sup>b</sup> |  | Training set<sup>c</sup> |  |
| --- | --- | --- | --- | --- | --- | --- |
|  | n | % | n | % | n | % |
| Patients | 6660 | - | 2412 | - | 5250 | - |
| Cases | - | - | - | - | 5341 | - |
| Samples | - | - | - | - | 5711 | - |
| GEX | - | - | - | - | 5857 | - |
| **OAS** |  |  |  |  |  |  |
| Median<sup>d</sup> (range<sup>d</sup>) Follow-Up Time Years | 7.0 (0.2-11.2) | - | 9.4 (1.1-11.2) | - | 5.8 (0.2-11.9) | - |
| Events | 1017 | 15.3 | 464 | 19.2 | 827 | 14.8 |
| Unknown | 3 |  | 0 |  | 286 |  |
| **DRFi** |  |  |  |  |  |  |
| Median<sup>d</sup> (range<sup>d</sup>) Follow Up Time years | 5.4 (0-10.9) | - | 8.1 (0.1-10.9) | - | 5.1 (0-11.1) | - |
| Events | 379 | 7.5 | 184 | 7.6 | 270 | 7.9 |
| Unknown | 1626 |  | 3 |  | 2444 |  |
| **Age at diagnosis** |  |  |  |  |  |  |
| Median (range) years | 65 (25-95) | - | 65 (25-95) | - | 65 (25-95) | - |
| ≤50 | 1373 | 20.6 | 499 | 20.7 | 1178 | 21.7 |
| >50 | 5287 | 79.4 | 1913 | 79.3 | 4255 | 78.3 |
| Unknown | 0 |  | 0 |  | 424 |  |
| **ER Status** |  |  |  |  |  |  |
| Positive | 5678 | 85.7 | 2073 | 86 | 4367 | 84.6 |
| Negative | 946 | 14.3 | 337 | 14 | 792 | 15.4 |
| Unknown | 36 |  | 2 |  | 698 |  |
| **PR Status** |  |  |  |  |  |  |
| Positive | 4714 | 71.2 | 1764 | 73.2 | 3568 | 69.2 |
| Negative | 1908 | 28.8 | 645 | 26.8 | 1588 | 30.8 |
| Unknown | 38 |  | 3 |  | 701 |  |
| **Node Status** |  |  |  |  |  |  |
| Positive | 2310 | 35.5 | 877 | 36.7 | 1800 | 36.1 |
| Negative | 4194 | 64.5 | 1514 | 63.3 | 3184 | 63.9 |
| Unknown | 156 |  | 21 |  | 873 |  |
| **HER2 Status** |  |  |  |  |  |  |
| Positive | 801 | 12.6 | 278 | 11.6 | 636 | 13.6 |
| Negative | 5571 | 87.4 | 2122 | 88.4 | 4051 | 86.4 |
| Unknown | 288 |  | 12 |  | 1170 |  |
| **Tumor Size (mm)** |  |  |  |  |  |  |
| Median (range) | 17 (0-250) | - | 20 (0-115) | - | 21 (0-250) | - |
| ≤10 mm | 952 | 14.6 | 311 | 13 | 738 | 15.1 |
| >10 ≤20 mm | 3382 | 51.8 | 1249 | 52.3 | 2445 | 50.2 |
| >20 mm | 2196 | 33.6 | 830 | 34.7 | 1689 | 34.7 |
| Unknown | 130 |  | 22 |  | 985 |  |
| **Grade** |  |  |  |  |  |  |
| 1 | 1026 | 16.1 | 369 | 15.7 | 752 | 15.9 |
| 2 | 3185 | 50.1 | 1159 | 49.2 | 2387 | 50.3 |
| 3 | 2152 | 33.8 | 829 | 35.2 | 1602 | 33.8 |
| Unknown | 297 |  | 55 |  | 1116 |  |
| **Histological type** |  |  |  |  |  |  |
| Ductal | 5177 | 78 | 1936 | 80.5 | 3950 | 76.6 |
| Lobular | 917 | 13.8 | 311 | 12.9 | 733 | 14.2 |
| Ductal/Lobular mixed | 128 | 1.9 | 32 | 1.3 | 120 | 2.3 |
| Other | 415 | 6.3 | 126 | 5.2 | 357 | 6.9 |
| Unknown | 23 |  | 7 |  | 697 |  |
| **Clinical Subgroup** |  |  |  |  |  |  |
| ER+/HER2-/N0 | 3197 | 50 | 1207 | 50.3 | 2279 | 47.7 |
| ER+/HER2-/N+ | 1727 | 27 | 666 | 27.8 | 1309 | 27.4 |
| HER2+/ER- | 254 | 4 | 85 | 3.5 | 221 | 4.6 |
| HER2+/ER+ | 564 | 8.8 | 193 | 8 | 462 | 9.7 |
| TNBC | 649 | 10.2 | 248 | 10.3 | 509 | 10.6 |
| Unknown | 269 |  | 13 |  | 1077 |  |
| **Clinical Subgroup** |  |  |  |  |  |  |
| ER+/HER2-/N0/Age>50 | 2663 | 41.3 | 1008 | 41.9 | 1890 | 38.3 |
| ER+/HER2-/N+/Age>50 | 1333 | 20.7 | 521 | 21.7 | 1001 | 20.3 |
| Other | 2456 | 38.1 | 877 | 36.5 | 2044 | 41.4 |
| Unknown | 208 |  | 6 |  | 922 |  |

<sup>a</sup>The early-stage follow-up cohort have no patient redundancy, i.e., contain one GEX profile per patient.
<sup>b</sup>The test set is a subset fully within the early-stage follow-up cohort. There is no overlap between the test set and training set.
<sup>c</sup>Part of the training set overlap with the early-stage follow-up cohort. Statistics for the training set are summarized on GEX.
<sup>d</sup>Median and range calculated for patients without event.
Percentage for variables with value exclude Unknown in total.

### Code availability statement

Derived SSP models will be available as functions in a standalone R package.

## Results

### Study cohorts

During the inclusion period, 11790 patients provided an informed SCAN-B consent based on either a diagnosis of BC or a suspected BC. In the current study, we included 7743 enrolled patients, based on availability of tissue and RNA-sequencing data, with a total of 8350 gene expression profiles (GEXs) generated from obtained tissue specimens as described ^22^ (including GEXs from bilateral diagnoses, multiple patient specimens, and repeated RNA-sequencing experiments). We assigned these patients to three partly overlapping cohorts as shown in Table 1 and Supplemental Figure 1a: i) a training set agnostic to variables other than available GEX in order to maximize training size for SSP models, i.e., including for instance multiple GEX profiles per patient and irrespective of verified BC or suspected BC diagnosis, ii) a test set with 2412 early-stage IBC patients, and iii) a larger cohort with 6660 IBC patients, referred to as the early-stage follow-up cohort hereon. As shown in Supplemental Figure 1a, the 6660-patient early-stage follow-up cohort overlapped with both the training set (partial overlap) and the test set, while the test set is completely independent from the training set. The early-stage follow-up cohort and hence the test set are non-redundant, i.e., included patients are represented by only one GEX profile. The reason for creation of the 6660-patient cohort (using clinical data obtained from the NKBC as outlined in Supplemental Figure 1b) was to form a cohort representative of the underlying IBC background population of the catchment area of the SCAN-B study in Sweden (Supplemental Figure 1c). Based on the early-stage follow-up cohort we could define a suitable independent test set for validation, selecting the majority of patients diagnosed between 2010-2013 in order to prioritize long follow-up time (median of 8.1 years for DRFi). In addition, the 6660-cohort also allowed us to naively assess the potential impact of SSP classification on treatment recommendations in a population representative manner. The usage of the different SCAN-B derived data subsets as well as external validation datasets is schematically shown in Figure 1a.

### Training SSP models for molecular subtypes of breast cancer

We trained an SSP model for five subtypes (SSP-PAM50) on the subtype classes assigned by our NCN model (NCN-PAM50) (Figure 1b). In total, 5255 GEX profiles were used in the training: Basal-like n=552, Her2-enriched n=528, Luminal A n=2573, Luminal B n=1377, and Normal-like n=225. The maximum overall agreement was observed at 24 gene-rules per subtype (Supplemental Table 1). Only marginal improvement was observed using >15 gene pair rules, consistent with previous reports ^14^. The number of unique genes (Entrez ID) represented in the selected model rules for all five subtypes was 216, of which 27 overlap with the reported PAM50 genes. The overall accuracy of SSP-PAM50 for predicting NCN-PAM50 in the training set was 85%.

An SSP model with four subtypes (SSP-Subtype) that would correspond to Prosigna subtypes was trained on NCN-Subtype labels (Figure 1b) from 5202 GEX profiles: Basal-like n=578, Her2-enriched n=529, Luminal A n=2718, and Luminal B n=1377. The maximum overall agreement in training was observed at 21 gene-rules per subtype (Supplemental Table 1), with only marginal improvement observed beyond 10 gene pair rules. The number of unique selected genes was 153, of which 27 genes overlap with PAM50 genes. The overall agreement of SSP-Subtype for predicting NCN-Subtype in the training set was 90%.

### Concordance between SSP and NCN for molecular subtypes in the independent test set

SSP models for molecular subtype were validated in our reserved test set of 2412 patients (Figure 1b). Overall agreement between SSP and NCN classifications for PAM50 (five subtypes) was 85% (Kappa=0.78) (Supplemental Figure 2a and Supplemental Table 2). The agreement is equivalent to the corresponding estimate from the training set, indicating that over-fitting has not occurred, and is higher than what was reported in the original AIMS study (77%) ^14^. The overall agreement for PAM50 (five subtypes) remains high (83%) even when 55 cases assigned as unclassified by NCN are regarded as discordant. For SSP-PAM50, the largest individual group of discordance is Luminal A by NCN assigned as Normal-like by SSP (128/1212 cases, 11%), consistent with findings in the original AIMS study ^14^. Other similarities with the original AIMS study include groups of Luminal B and Normal-like by NCN assigned as Her2-enriched and Luminal A respectively by SSP (Supplemental Figure 2a). However, for most groups of discordant assignments, their respective fraction of the NCN defined subtype is low by comparison. Overall agreement between SSP and the original AIMS method for PAM50 subtype in our validation cohort was 74% (Kappa=0.63) and corresponding overall agreement between NCN and AIMS was 70% (Kappa=0.56). The majority of the discordance occurred between Luminal A by AIMS vs. Luminal B by SSP (41% of discordant cases) and Normal-like by AIMS vs. Luminal A by SSP (36% of discordant cases).

The agreement between SSP and NCN for Subtype (four subtypes) in the test set was 90% (Kappa=0.84) (Supplemental Figure 2b and Supplemental Table 2). Here, the largest group of discordance in absolute numbers was 52 of 1311 (4%) Luminal A by NCN assigned as Luminal B by SSP, followed by 46 of 582 (8%) Luminal B by NCN assigned as Her2-enriched by SSP (Supplemental Figure 2b). No individual group of discordant subtype assignment by SSP represented >8% of the NCN defined subtype.

### Training and validation of an SSP model for ROR in breast cancer

Since ROR score is an integer value between 0-100 we used data binning with 20 equally spaced levels to transform NCN-ROR into categorical training labels for our SSP-ROR model (Supplemental methods). For training the SSP-ROR model, NCN-ROR scores from a total of 5359 GEX profiles were stratified into binned ROR labels: <5 n=100, 6-10 n=186, 11-15 n=272, 16-20 n=314, 21-25 n=324, 26-30 n=317, 31-35 n=361, 36-40 n=318, 41-45 n=311, 46-50 n=364, 51-55 n=384, 56-60 n=390, 61-65 n=411, 66-70 n=366, 71-75 n=349, 76-80 n=272, 81-85 n=196, 86-90 n=124 profiles. The maximum overall agreement in training was observed at 21 gene-rules per subtype (Supplemental Table 1) with only marginal improvements observed using >10 gene pair rules. The union of unique genes represented in selected rules for all ROR bins was 296, of which 18 overlapping with the PAM50 genes. The overall accuracy in the training set for categorical binned ROR labels was 17%. A strong linear relationship was observed between the SSP and NCN categorical value (R^2^=0.87).

In the test set, overall agreement for binned NCN-ROR score and predicted SSP-ROR was 17% (Kappa=0.13, weighted Kappa=0.90), equivalent to training results. Similar to the training set, a strong linear relationship with binned ROR score (R^2^=0.88) was also observed in the test set (Supplemental Figure 2c). The relationship was also visualized by boxplots of the non-binned NCN-ROR scores for SSP-ROR (Supplemental Figure 2d). Importantly, when stratified by SSP-PAM50, the distributions of ROR showed similar relationships between subtypes for NCN-ROR scores (Supplemental Figure 2e) and SSP-ROR score (Supplemental Figure 2f). Also, the distributions were as expected lower in Luminal A cases ^7,8^. Distributions of ROR scores stratified by SSP-Subtype were also specifically investigated for Luminal A and Luminal B cases within the clinical subgroup of ER+/ HER2-tumors (Supplemental Figure 2g-h), again finding relationships between subtypes to be similar and also consistent with what has been reported for the Prosigna assay (^8^ and Prosigna insert 2017-07 LBL-C0191-09 section 15.1 Figure 9).

SSP and NCN concordance for ROR score was also investigated after relevant stratification into ROR risk category groups (Low, Intermediate, High) using cutoffs specific for nodal status ^3^ and used by Prosigna (^8,43^ and Prosigna insert 2017-07 LBL-C0191-09 section 13.4, Table 9). For NCN-ROR, the score was calculated using the gross tumor size variable as described ^3^. For SSP-ROR, the assigned score was adjusted with +5 for tumors >20mm to appropriately account for the effect of the gross tumor size variable and minimizing the risk of underestimating the score (Supplemental methods). Overall agreement between SSP and NCN for risk category was 84% (Kappa=0.75, weighted Kappa=0.90) (Supplemental Figure 2i). Effectively, all discordance (>99%) was observed between adjacent risk groups, explaining the high weighted Kappa, with discordance between Intermediate by NCN-ROR and either Low or High classification by SSP-ROR, comprising 40% and 33% of all discordant cases respectively. To illustrate and evaluate agreement that reflects practical clinical use we also performed a dichotomized comparison by combining Low and Intermediate risk classification into one category, similar to the study by Bartlett et al. ^5^ that compared different commercial multigene tests in BC. The overall agreement between SSP and NCN for this two-group ROR stratification was 92% (Kappa=0.84) (Supplemental Figure 2j).

### Training and validation of SSP models for clinical markers in breast cancer

In addition to deriving SSPs for intrinsic molecular subtypes and ROR score, we also trained SSP models for five conventional clinical BC markers, ER, PR, HER2, Ki67, and NHG, using training labels based on clinicopathological registry data (Figure 1c). Cutoffs for ER and PR status were set to 10% or greater positive staining according to Swedish national guidelines. For HER2, in addition to a general SSP model, we also trained two separate SSP models specific for ER status using only ER+ or ER-cases respectively. For Ki67 status, a two-group model (High/Low) was trained using cutoffs for included tumors from the respective local pathology departments. The number of gene-rules per class at maximum overall agreement in training ranged from 3 for PR to 19 for ER (Supplemental Table 3). Performance was first evaluated in the independent test set (Table 2 and Supplemental Figure 3). Concordance with clinicopathological status was very high for ER (overall accuracy=96%, Kappa=0.86) and high for PR (overall accuracy=87%, Kappa=0.70) with clinically relevant positive predictive values (99% and 95%, respectively). Concordance was high for HER2 using ER-specific models (overall accuracy=92%, Kappa=0.67) and moderate for the general HER2 SSP model (overall accuracy=89%, Kappa=0.58). Overall concordance for HER2 SSP models was mainly negatively influenced by false positives for the SSP models. Correspondingly, the negative predictive values were high (98% for both the general and ER specific HER2 models). Concordance with clinicopathological status was moderate for Ki67 (overall accuracy=80%, Kappa=0.59), and fair for NHG (overall accuracy=57%, Kappa=0.38, weighted Kappa=0.60). For NHG, 80% of all discordance was from NHG Grade 2 (811 of in total 1012 discordant cases), stratified by SSP into Grade 1 or 3. This observation is consistent with previous studies dividing NHG Grade 2 tumors into low and high-proliferative cases ^44,45^. By comparison, only a small fraction of discordant cases (58/1012, 5.7%) were misclassified from Grade 1 to 3 or vice versa reflected by substantially higher weighted agreement bordering moderate and high concordance. The negative predictive values were accordingly comparably high for both Grade 1 and Grade 3 (95% and 90%, respectively). To test whether discordant SSP stratification of clinical NHG status provided prognostic value we created Kaplan-Meier plots in the subgroup of patients with ER+/HER2-disease who only received endocrine adjuvant treatment (Supplemental Figure 4). A marked difference in DRFi was found for stratification of clinical NHG Grade 2 but not of clinical NHG Grade 1 or 3 (Supplemental Figure 4).

**Table 2.**
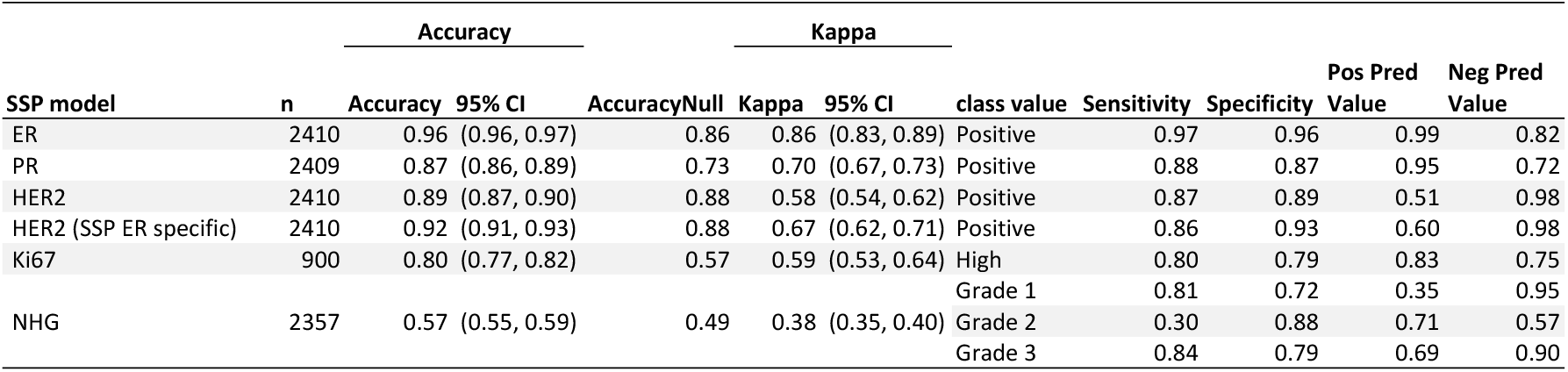
SSP performance validated against clinicopathological registry data (NKBC) in the independent population-based SCAN-B test set.

The quality of registry data in NKBC has been shown to be high ^46^. In addition, review of medical chart data performed in subsets of the SCAN-B cohort has shown high validity of register data in general and for dichotomized treatment data in particular, e.g., yes/ no for endocrine and chemotherapy treatment ^27^. Even so, to further investigate if evaluated concordance for clinical markers was adversely affected by assessment against registry data we evaluated performance against consensus status from re-stains and re-evaluation done by three board-certified breast cancer pathologists in the independent ABiM material ^23^. Concordance was largely comparable with results in the SCAN-B test set. For Ki67, distributions appeared skewed using the Ki67 cut-off set at High >20%, as a large number of cases were given a re-evaluated consensus score of exactly 20% and most of these were classified as High by SSP (Supplemental Table 4 and Supplemental Figure 5).

### Comparison between NCN and SSP stratifications by patient outcome

To further validate SSP subtype and ROR models against NCN stratification on a group level, we assessed prognostic value by survival analysis using registry data in the population representative test set (Figure 1b). Comparison with patient outcome is particularly relevant as it reflects the intended use of the classifications. Moreover, we reasoned that group level comparisons are relevant given the nature of the intrinsic subtypes and ROR classification with classes defined by underlying boundaries for relative correlations to centroids. As such, there are no obvious distinctions in underlying data between some classes (e.g., Luminal A vs B, or a continuous ROR score).

Outcome analysis typically requires comparisons within groups of uniformly treated patients. However, as the test set represents early-stage IBC in Sweden, the majority diagnosed between 2010-2013 and all treated in accordance with national guidelines at the time of study inclusion, the overall differences in outcome for intrinsic subtypes can be expected to reflect treatment outcome for each respective group. Therefore, we first compared outcome characteristics for molecular subtypes by Kaplan-Meier plots using DRFi in the full test set and irrespectively of clinical markers and administered treatment (Figures 2a-b). As expected, intrinsic subtypes display markedly separate outcome consistent with previous reports ^7,47^. Patient outcomes are generally very good and highly similar between SSP and NCN with respect to Luminal A and Luminal B cases, with each respective group having 95% (Luminal A) and ∼85% (Luminal B) event-free survival respectively irrespective of classification method and model.

**Figure 2.**
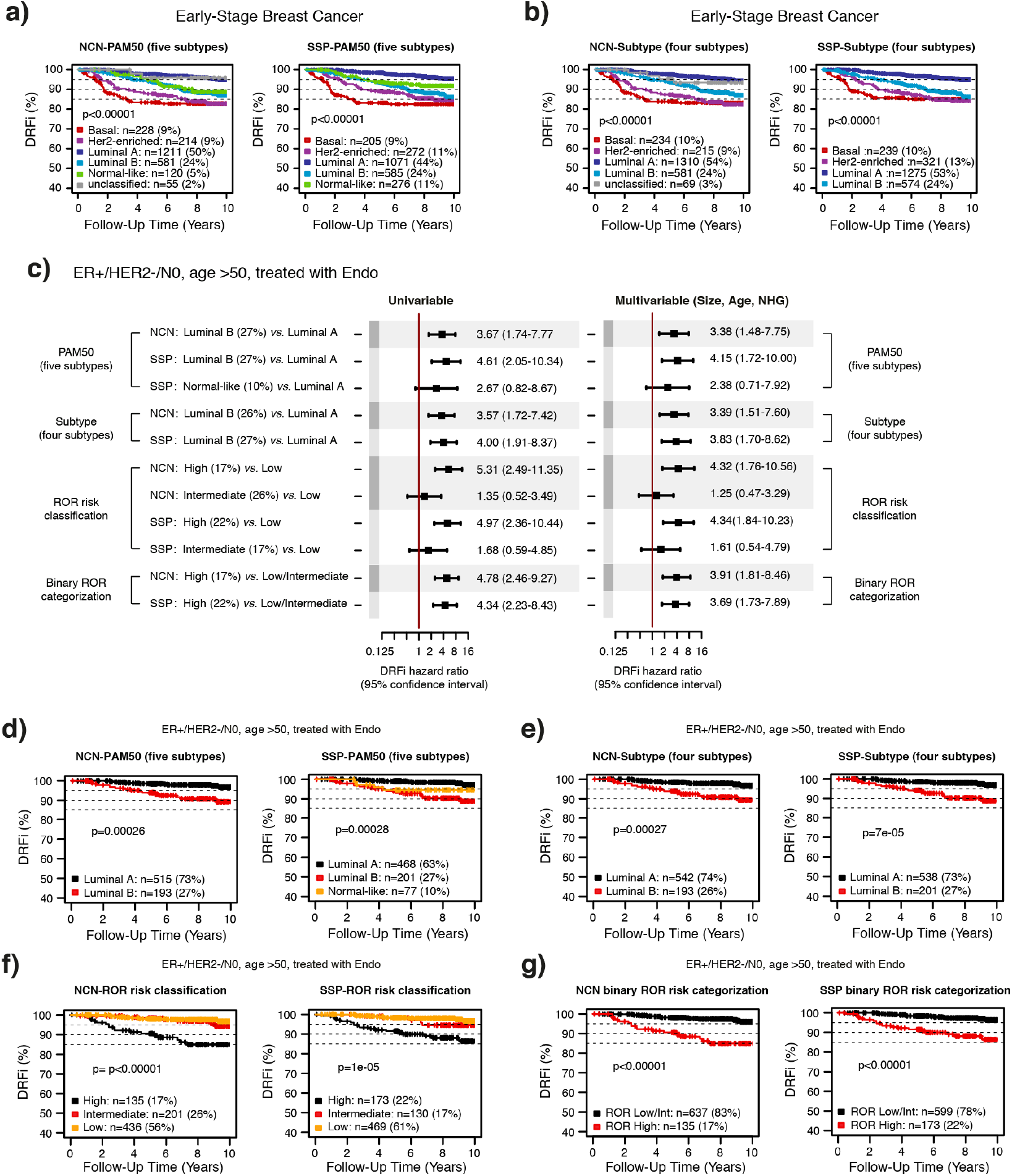
Assessment of prognostic value of SSP stratification and NCN stratification. Comparison of SSP and NCN classifications in the independent population-based test set by assessment of prognostic value. Kaplan-Maier plots for molecular subtype with five groups (PAM50) or four groups (Subtype) using DRFi as clinical endpoint: **(a)** PAM50 by NCN (left) and SSP (right), **(b)** Subtype by NCN (left) and SSP (right). **(c)** Cox regression analysis using DRFi as endpoint in the test set restricted to patients with ER+/ HER2-/N0 disease diagnosed over 50 years of age that only received endocrine adjuvant treatment (n=772). Test and reference group is specified on the left. Hazard ratios and 95% confidence interval ranges from univariable analysis (left forest plot) and multivariable analysis (right forest plot) with tumor size, age at diagnosis, and NHG as covariates. Kaplan-Meier plots for stratification of ER+/ HER2-/N0 disease diagnosed over 50 that only received endocrine adjuvant treatment in the test set by NCN (left in each panel) and SSP (right in each panel) for: **(d)** PAM50 subtype, **(e)** Subtype, **(f)** ROR risk classification, and **(g)** and the two-group ROR stratification according to Bartlett et al. ^5^.

We next focused on specific patient subgroups where the available commercial molecular assays in question are generally recommended for use in assisting treatment decisions. Prognostic value was first assessed for NCN and SSP-based intrinsic subtypes and for different ROR classification groups in patients >50 years with ER+/HER2-/N0 disease that only received endocrine adjuvant treatment (n=772). Hazard ratios and 95% confidence intervals were highly similar and overlapping between corresponding SSP and NCN based stratifications in univariable analysis (Figure 2c, left side), as well as in multivariable analysis with tumor size, age at diagnosis, and NHG as covariates (Figure 2c, right side). To further illustrate SSP and NCN stratifications we generated Kaplan-Meier plots showing similar DRFi characteristics for stratifications by SSP and NCN for PAM50 subtype (Figure 2d), Subtype (four subtypes) (Figure 2e), ROR risk group classification (Figure 2f), and the two-group stratification according to Bartlett et al. ^5^ (Figure 2g).

The same tendency for outcome and similarity between SSP and NCN stratifications was also observed in patients with ER+/HER2-/N0 tumors that received adjuvant chemotherapy prior to endocrine treatment, although groups are small as shown for ROR risk classification (Supplemental Figures 6a-b). Furthermore, comparable prognostic stratification for SSP and NCN was seen in the similarly sized group of patients with ER+/ HER2-/N0 tumors that received no adjuvant treatment (Supplemental Figures 6c-d). Here, the majority of tumors were classified as low risk by ROR score and reassuringly with none or very few distant recurrences. Finally, the same pattern also extended to ROR stratification of node positive (N+) disease and the larger group of patients with ER+/HER2-/N+ tumors that only received adjuvant endocrine treatment (Supplemental Figures 6e-f). Within this group, about 50% of cases were categorized as either Low or Intermediate risk by ROR and contribute few events.

### Cross-comparison between SSP and Prosigna in independent external clinical series

To benchmark the developed SSP models for Subtype and ROR versus a commercially available assay, we compared classifications to results from the Prosigna assay performed on FFPE tissue in two independent external clinical series (Figure 1b): i) OSLO2-EMIT0 (n=103, clinical Prosigna assay results), and ii) ABiM (n=100, Prosigna classifications calculated from non-clinical Nanostring data). Overall accuracy for Subtype assignment was 83% in OSLO2-EMIT0 and 80% in ABiM (Kappa=0.73 and 0.72, respectively) (Table 3, Supplemental Figure 7a and Supplemental Figure 8a). Overall accuracy for ROR risk category was 68% in OSLO2-EMIT0 and 84% in ABiM (Kappa=0.50 and 0.70, weighted Kappa=0.70 and 0.78, respectively). We also compared distributions of ROR score by subtype assignment in the respective full series as well as restricted to ER+/HER2-tumors classified as Luminal subtype (Supplemental Figure 7d-g and Supplemental Figure 8d-g). Concordance for two-group stratification reported by Bartlett et al. ^5^ was 82% in OSLO2-EMIT0 and 89% in ABiM (Kappa=0.64 and 0.76, respectively) (Table 3, Supplemental Figure 7i and Supplemental Figure 8i). In a pooled analysis of OSLO2-EMIT0 and ABiM the agreement for Subtype was 81% (Kappa=0.73), 76% for ROR risk category (Kappa=0.59, weighted Kappa=0.74), and 85% for the Bartlett two-group stratification of ROR risk (Kappa=0.70). In a pooled analysis restricted to ER+/HER2-tumors agreement for Subtype was 79% (Kappa=0.61), 75% for ROR risk (Kappa=0.61, weighted Kappa=0.74), and 86% for the Bartlett two-group stratification of ROR risk (Kappa=0.72).

**Table 3.**
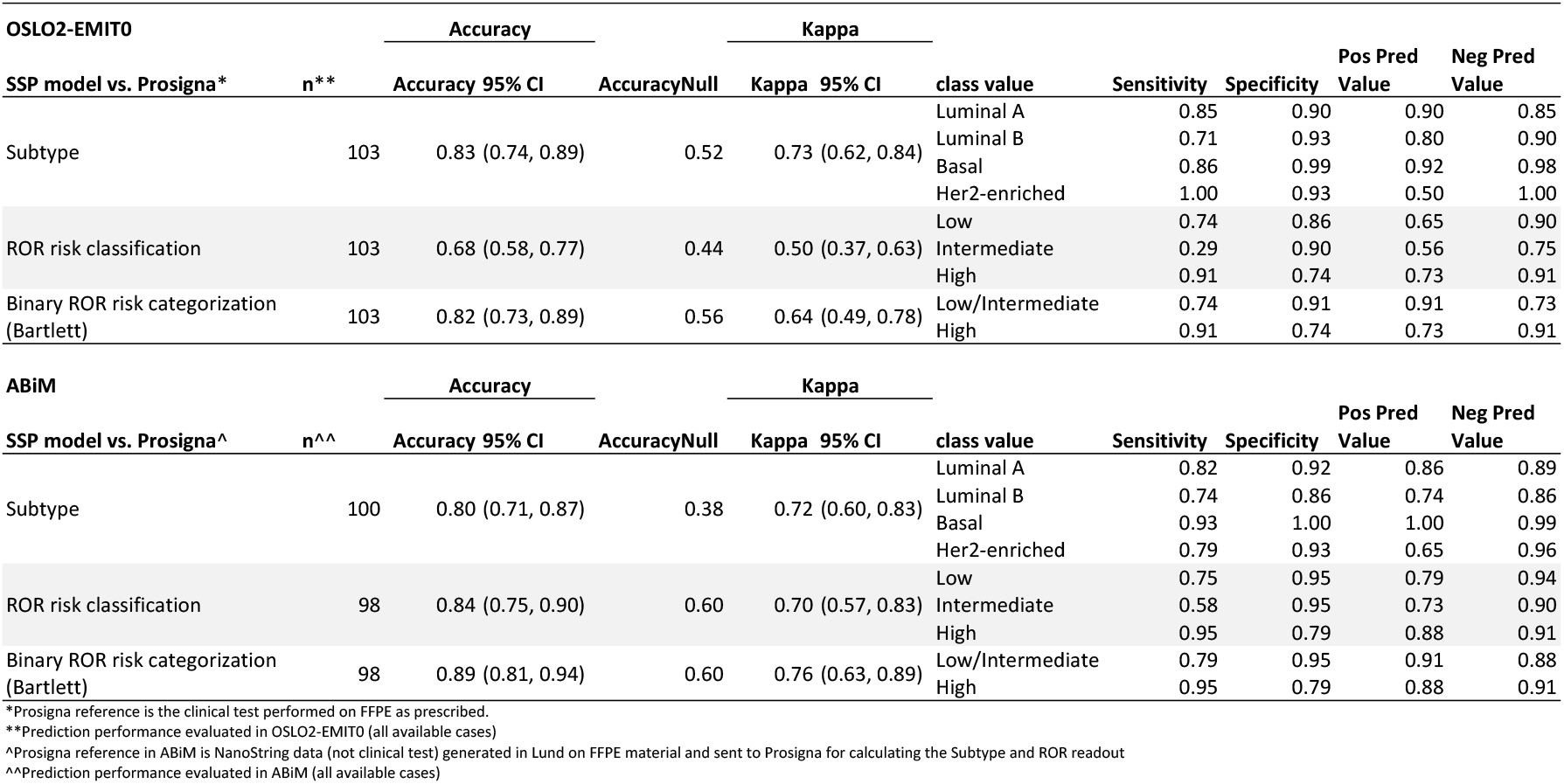
SSP prediction performance validated against Prosigna classification in the full external clinical series OSLO2-EMIT0 and ABiM.

Expected confounders in the above comparisons would include sampling and tissue preservation (FFPE versus fresh-frozen) as noted by others for the clinical Prosigna assay ^40,43^. To assess these confounders and put concordances in a context we utilized multiple readings from different models and procurements in the ABiM cohort (Figure 1b). We first compared concordance between SSP and Prosigna using classifications from applying the respective models to data from the same RNA extract obtained from macro-dissected fresh tissue. Overall agreement for Subtype assignment increased to 87% (Kappa=0.81), in line with agreement between SSP and NCN models in the test set. Notably, ROR risk classification decreased to 79% (Kappa=0.61, weighted Kappa=0.76), whereas concordance for the Bartlett two-group risk category remained unchanged at 89% (Kappa=0.76) (Supplemental Table 5 top section). Finally, to conversely isolate the effect of sampling on discordance we evaluated agreement in the ABiM cohort solely using the Prosigna model but comparing data obtained from either FFPE tissue or from the paired macro-dissected fresh tissue. The overall agreement for Subtype assignments was 82% (Kappa 0.74), for ROR risk classification 84% (Kappa 0.71, weighted Kappa=0.84) and 90% (Kappa 0.79) with two-group categorization (Supplemental Table 5 bottom section), largely matching the agreements between SSP and Prosigna.

For a more specific comparison between SSP and Prosigna that better reflects the use for directing adjuvant chemotherapy in a clinical setting, we compared agreement for stratification that emulates treatment recommendation (ETR) in postmenopausal patients with ER+/HER2-/N0 and pT1-2 tumors adapted from Norwegian national guidelines (Supplemental methods). The ETR schema adheres to actual general recommendations but does not include individualized assessment of possible escalation or de-escalation. The treatment guidelines stratify patients into three groups with respect to recommended adjuvant treatment: None, Endo (i.e., endocrine treatment alone for 5-10 years), or ChemoEndo (i.e., adjuvant chemotherapy followed by endocrine treatment for 5-10 years). The comparison was also done for treatment recommendations dichotomized for chemotherapy, i.e., by combining None and Endo into one group. SSP performance was evaluated in both the pooled external clinical series (n=87) as well as in the respective series separately (Supplemental Table 6). Overall agreement for ETR in the pooled data was 78% (Kappa=0.65) and when dichotomized for chemotherapy was 85% (Kappa=0.66).

### Assessing potential impact of SSP molecular subtype and ROR testing on use of chemotherapy

In addition to validating SSP classifications for Subtype and ROR against research-based NC classifications and benchmarking SSP against Prosigna models we also wanted to apply our SSP models in the entire 6660-patient follow-up cohort (Figure 1, Table 1, Supplemental Figure 1) to assess the potential extent and type of altered treatment recommendation from using SSP models in treatment guidance. To this end we used the naïve ETR classification dichotomized for chemotherapy and compared this with information from NKBC on administered treatment.

To verify that SSP classifications were independent of year of diagnosis we first calculated the proportions of SSP-Subtype and SSP-ROR risk group in the entire population stratified by year of diagnosis. Proportions for classifications varied slightly between years but were largely stable throughout the enrollment period that extends over nine years (Figures 3a-b). Overall proportions across the entire period for SSP-Subtype were: 52% Luminal A, 25% Luminal B, 10% Basal-like and 13% Her2-enriched (Figure 3a). Corresponding proportions for SSP-ROR risk classification were: 38% Low, 15% Intermediate and 47% High (Figure 3b).

**Figure 3.**
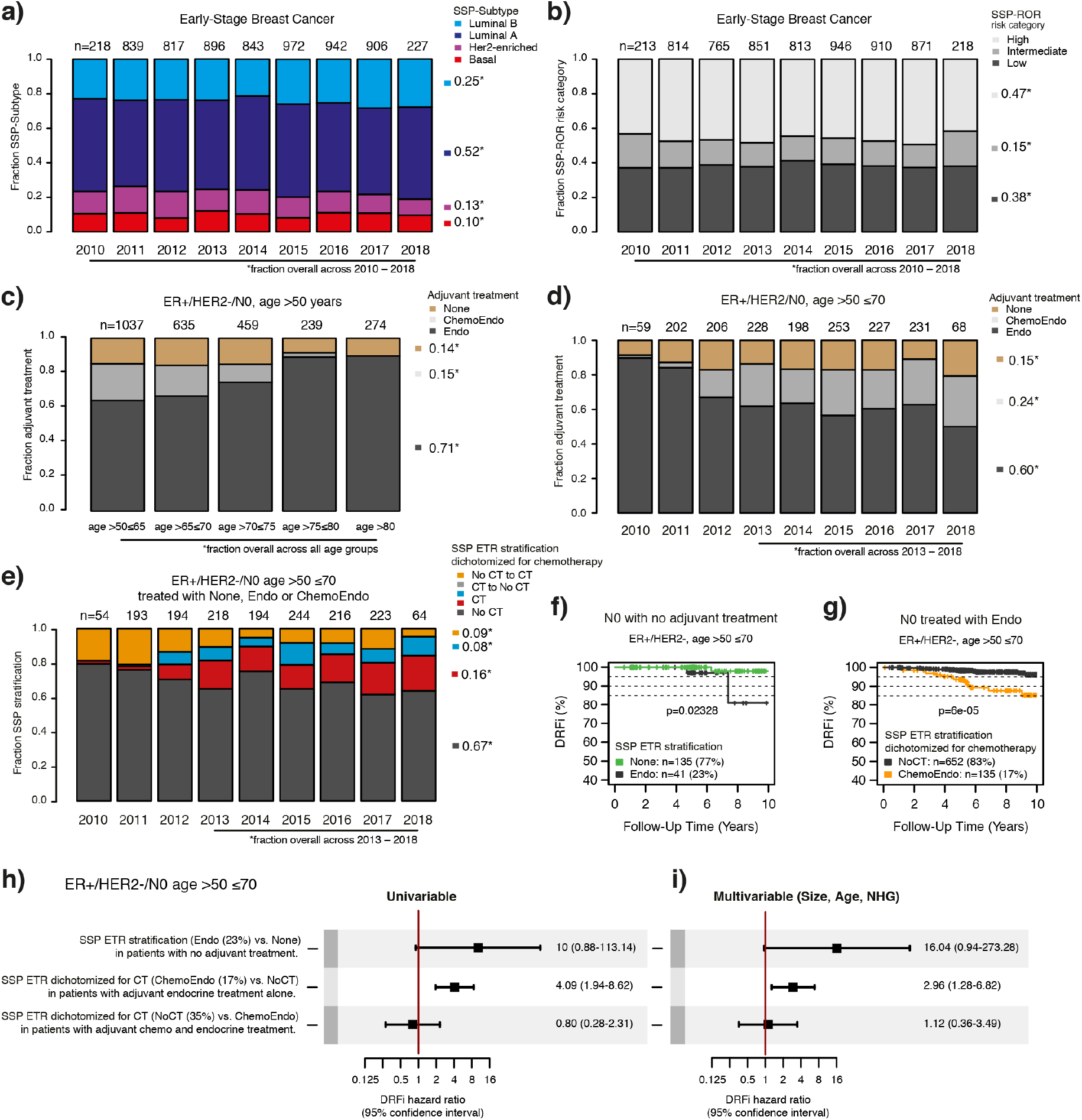
SSP classifications for Subtype and ROR risk category in early-stage breast cancer and cross-comparison with administered systemic treatment. The basis for comparisons is the 6660-patient follow-up cohort (Table 1). Summarized proportions are shown on the right side of bar graphs. The first and last year of enrollment (2010 and 2018) are not full calendar years and therefore include notably smaller numbers of enrolled patients. **(a)** Proportions of SSP-Subtype by year of diagnosis. **(b)** Proportions of SSP-ROR risk category by year of diagnosis. **(c)** Proportions for adjuvant treatment within ER+/HER2-/N0 patients diagnosed at age >50 years by different age at diagnosis. Endo: endocrine therapy only, ChemoEndo: adjuvant chemotherapy and endocrine therapy. None: no adjuvant systemic therapy. **(d)** Proportions for adjuvant treatment within ER+/HER2-/N0 patients diagnosed at age >50 ≤70 years by year of diagnosis. **(e)** Cross-comparison of the naïve SSP ETR dichotomized for chemotherapy (yes/no) with records of administered systemic treatment within ER+/HER2-/N0 patients at age >50 ≤70. The groups for which SSP treatment recommendation is in agreement with the administered treatment are shown in black for regimen without chemotherapy (No CT) and in red for regimen including chemotherapy (CT). The discordant groups where SSP would lead to escalation of treatment (No CT to CT) are shown in orange and de-escalation of treatment (CT to No CT) in blue. **(f)** Kaplan-Maier plot for SSP stratification by SSP-ETR treatment recommendation within the N0 subgroup of ER+/HER2-patients diagnosed at age >50 ≤70 and no adjuvant treatment. **(g)** Kaplan-Maier plot for SSP stratification by SSP-ETR dichotomized for chemotherapy (chemotherapy vs. no chemotherapy) within ER+/HER2-/N0 patients diagnosed at age >50 ≤70 treated with adjuvant endocrine therapy only. **(h)** Forest plots of Hazard ratios and 95% confidence interval ranges from univariable and **(i)** multivariable Cox regression using DRFi as endpoint stratified using SSP treatment recommendation. Multivariable analysis is with tumor size, age at diagnosis, and NHG as covariates. Test (SSP stratification) and subgroup (administered treatment) is specified on the left of the univariable forest plot.

Among other clinical management indications, molecular testing is indicated for postmenopausal IBC patients with ER+/HER2-/N0 with an ambiguous risk of recurrence. To attempt to represent a relevant indication we first studied patients with known treatment status diagnosed with ER+/HER2-/N0 breast cancer at age >50 years (2644/6660, 40%). In this subgroup, the use of chemotherapy differed between age groups and adjuvant therapy among the more elderly patients was largely restricted to endocrine treatment (Figure 3c). Therefore, the final clinical assessment subgroup was restricted to patients aged >50 ≤70 years to reduce the impact of high age and expected associated comorbidities as factors influencing treatment decisions. The fraction of patients receiving chemotherapy in this age-restricted subgroup increases across early years of enrollment, especially apparent from 2012, but levels out for the later years (Figure 3d). The observed increase coincides with changes in national treatment guidelines introduced during the period of enrollment. Therefore, to better extrapolate our results, estimates were calculated for patients from the later enrollment period (2013-2018). Proportions of treatment across this latter period differ to some extent from the overall and were 60% (versus 65%) with endocrine only and 24% (versus 20%) with adjuvant chemotherapy and endocrine treatment, whereas the proportion of patient that received no adjuvant treatment remained at 15% (Figure 3d). The potential effect on therapy was estimated by cross comparing the naïve ETR dichotomized for chemotherapy with NKBC records of administered systemic treatment.

In the N0 subgroup strict adherence to ETR would result in modest net increased use of adjuvant chemotherapy from 24% to 25% estimated for patients from 2013-2018 (Figure 3e). The estimated net change is the combined result from patients where treatment would be escalated with chemotherapy (No CT to CT, 9%), and patients that would be spared chemotherapy (CT to No CT, 8%). Thus, in total 17% of the investigated clinical subgroup had potential for changed chemotherapy recommendation based on SSP molecular subtyping.

In addition to estimating possible effects on therapy, we also assessed the prognostic value of the molecular test by stratification of uniformly treated subgroups. For N0 patients with no adjuvant treatment, patients suggested for endocrine treatment by SSP (41 of 176) had worse outcome (Figure 3f) with a univariable Cox hazard ratio of 10 (95% CI=0.88–113) (Figure 3h) and multivariable hazard ratio of 16 (95% CI=0.94–273) (Figure 3i). For N0 patients with endocrine therapy only, patients suggested for escalation (No CT to CT) by SSP (135 of 787) had worse outcome (Figure 3g) with a univariable Cox hazard ratio of 4.09 (95% CI=1.94–8.62) (Figure 3h) and multivariable hazard ratio of 2.96 (95% CI=1.28–6.82) (Figure 3i). For N0 patients receiving adjuvant chemotherapy there was no difference in outcome for the patients suggested for de-escalation of chemotherapy by SSP classification (Figures 3h-i).

## Discussion

In this study we have trained, validated, and benchmarked RNA-sequencing based gene expression SSP models for conventional clinical markers, molecular subtypes, and ROR in the largest, consecutive, primary BC cohort reported worldwide to date. Importantly, the observational population-based SCAN-B cohort is representative of contemporary stage-distribution and treatments, with sampling of fresh tumor tissue completely integrated in parallel with clinical routines ^20,22^ and with a complete turn-around assay time compliant with clinical usage (see Supplemental methods). These characteristics strongly support that results based on this cohort can be extended and generalized to the national BC population in Sweden, and other populations with comparable demographics.

The SSP classifications for conventional markers were validated against clinical pathology data from NKBC and against consensus status from independent re-assessment by three pathologists in the ABiM cohort ^23^ (Figure 1c). The concordance was high for ER, PR and HER2. For ER and PR results are well in line with previous studies confirming that sequencing-based assays can accurately mimic readouts from current commercial assays ^23,48,49^. However, for any marker that is a direct target for treatment, such as the ER and HER2 receptors, special considerations regarding practical use of surrogate assays are required. For example, for HER2, the negative predictive value was very high for all SSP models (NPV=0.98). Considering that the Swedish *HER2* amplification rate in 2019 was 13.5% ^25^ this would imply that the sequencing-based classifiers could drastically reduce the number of negative tests performed. The lower positive predictive value of HER2 could be explained by differences in assessments. While histochemical or *in situ* hybridization scoring takes only stained invasive cancer cells into account, gene expression is measured from bulk RNA extracts including intraductal components. Moreover, the SSP models may also capture elevated HER2 signals present in tumors without HER2 protein overexpression or gene amplification. This suggests that a positive SSP HER2 scoring should be complemented with *in situ* (FFPE HER2 IHC/ISH) measurements to assure a correct status for anti-HER2 treatment decisions. On the other hand, tumors with elevated HER2 signal but without protein overexpression or gene amplification may potentially be sensitive to other treatments targeting the HER2 signaling pathway, which should be further addressed. In contrast to ER, PR and HER2, the SSP classifications for the proliferation marker Ki67 and NHG had lower accuracy. This may be expected as these conventional markers are routinely assessed by means that have been reported as sensitive to subjective and inter-observer variability ^23,50-52^. Perhaps more importantly, the sequencing-based predictor showed very high negative predictive values for NHG Grade 1 and 3 tumors, while for the more heterogeneous NHG Grade 2 tumors the sequencing-based classification showed the ability to stratify patients into subsets with different clinical outcome, representing actual clinical value, in line with previous studies in the field ^23,44,45^.

A primary focus of this study was to derive SSP models for molecular subtypes and ROR scores in BC that would closely mimic conventional nearest centroid classifications, while being more applicable for inclusion in routine diagnostics of early BC. To achieve this, we used a meticulous normalization approach to generate suitable centroid training labels, followed by conventional class agreement analysis, patient outcome analysis, benchmarking versus Prosigna, and assessment of the impact on treatment recommendation (Figure 1b). Consistently, concordance evaluated in the independent test was high to very high (85-90%) between SSP subtype models (SSP-PAM50 and SSP-Subtype) and corresponding NCN classifications. Moreover, the main classification discrepancies were observed at the expected boundaries between the Luminal A and Luminal B subtypes, and between the Luminal A and Normal-like subtypes. This is not surprising given the underlying definition for discriminating between the subtypes, for which the distinction between Luminal A and B is determined by the ratio of relative correlations to the respective centroids. In reality there is no distinct separation in the underlying data discriminating between these two subtypes, rather, they are two ends of the same spectrum of relative ratios. Thus, cases in the middle of the spectrum are no more Luminal A than Luminal B or vice versa, similar to the seamless transition between varying degrees of some biological processes such as cell proliferation ^53,54^. The same reasoning applies with more clarity to the ROR risk classification where original cut-points in ROR score between classes are set to achieve chosen incidences of disease recurrence. Although the coefficients for ROR score are derived to model risk of recurrence, it is still an association to risk of recurrence, i.e. a clinical endpoint. As such, there are no distinct underlying transitions between degrees of ROR score. Nonetheless, high agreement between SSP and NCN classifications were observed for ROR risk classifications as well as for a two-group comparison combining ROR Low and Intermediate risk classification into one category (Supplemental Table 2). Importantly, the high agreement was also mirrored on a group level by similar prognostic performance for SSP and NCN models when assessed in the independent test data (Figure 2), particularly within the relevant clinical subgroup of post-menopausal (>50 years) ER+/HER2-/N0 BC treated with endocrine treatment alone (Figures 2c-g). These analyses suggest that SSP and NCN models are exchangeable on a group level concerning prognostic value. Moreover, our results demonstrate that SSP models are on their own capable of further stratifying current clinical subgroups of BC into subgroups with different clinical outcome, representing potential real clinical value.

An important aspect of the current study compared to existing academic studies in the field is the benchmarking of SSP classifications to actual matched clinical Prosigna classification, or Prosigna classification based on non-clinical Nanostring data, in two external clinical cohorts. Despite the external series being comparatively small, the comparisons provide important insight and benchmarking against results from an available and validated assay that is in clinical use today. These direct class comparisons demonstrated moderate to high numerical agreements and broadly high numerical agreements in pooled analysis. When interpreting the benchmarking results, tissue heterogeneity and sampling procedure need to be acknowledged as potential sources of discordance. This was also highlighted by Nielsen et al. for the clinical Prosigna assay, reporting 90% and 93% average agreement of ROR risk category for N0 and N+ patients, respectively, based on analysis at different laboratory sites using different tissue sections from the same tissue blocks ^43^. Whereas Prosigna prescribes input material from FFPE sections verified to comprise a minimum of 10% invasive component, the RNA-sequencing based SSPs uses input material from fresh macro-dissected tumor tissue. This prevents strict direct comparisons of models even when samples originate from the same tumor. To investigate this issue, we also compared results from the SSP and Prosigna models with measurements from identically sourced RNA aliquots from fresh macro-dissected tissue. Notably, this improved the classification agreement for Subtype, decreased agreement for ROR risk category, while agreement for binary ROR risk classification remained the same. Importantly, performance for Prosigna from macroscopically evaluated fresh tissue against Prosigna from FFPE was in line with the corresponding agreements between SSP and Prosigna on FFPE. There are some weaknesses in these comparisons such as small sample size and diverging from Prosigna prescribed protocols; nonetheless, they highlight the soft transitions between classes and that exact agreement is neither expected nor needed for equivalence in practice as suggested by Bartlett et al.^5^.

Although the SSP vs. Prosigna classification agreement was not perfect, the concordance was high compared to reported agreement levels for different multigene assays ^21,55,56^, extending to clinical tests individually approved for the same use ^5^. Notably, in those studies different multigene assays showed far from perfect agreement on an individual sample basis. In contrast, up to 89% direct agreement between SSP and Prosigna as observed for binary ROR risk category in this study infers not only similar group level characteristics, but also high agreement to Prosigna results on the sample level, although continued confirmation is needed as larger cohorts with data for both assays becomes available.

Concluding that SSP classifications are at the least comparable to Prosigna classification on a population level, we next examined the naïve impact on treatment decisions had SSP results been available to guide recommendation of adjuvant chemotherapy. Acknowledging that guidelines have changed during the years of patient recruitment, the estimations presented here remain somewhat uncertain. Moreover, in clinical practice the molecular classification would only be one part of the decision process as multidisciplinary teams, when recommending treatment, also consider co-morbidity and patient preferences. Such factors have not been accounted for in our analysis even though we restricted the assessment groups to age 51-70 years to reduce the effect of old age and accompanying co-morbidity. Moreover, we compared emulated treatment considerations with registry data for administered treatment, thus our estimates remain to be confirmed by future prospective evaluation. Nonetheless, our assessments suggest that naïve usage of SSP recommendations for treatment decisions would lead to a modest net change in use of adjuvant chemotherapy for ER+/HER2-/N0 BC in Sweden. This contrasts an observation in a North American population of early-stage BC, where a 70-gene signature identified a significant proportion of clinically high-risk patients that might not require chemotherapy ^2^. However, hitherto presented data on change of therapy for the Prosigna test is more in line with our results ^57^. Still, our results highlight that within ER+/HER2-/N0 early-stage disease diagnosed at >50≤70 years as much as 17% of cases may be subjects for changed chemotherapy recommendations as a result of molecular subtyping. In this study we did not include the corresponding assessment in node positive disease, as current national guidelines do not include these. However, it seems likely that a similar or even larger fraction of cases in this group would be subjects for changed treatment recommendations as a result of molecular subtyping.

The clinical usefulness of currently available and validated multigene assays is limited to certain subgroups of BC patients. For instance, in Sweden the recently added recommendation in the national guidelines ^25^ to use molecular diagnostics was limited to early-stage postmenopausal ER+/HER2-/N0 BC. Moreover, current clinical gene expression-based assays are associated with a substantial financial cost, may require sending samples outside a regional healthcare region, and typically report a single assay output, e.g., a treatment recommendation for a specific patient population. Considering that modern cancer diagnostics require a multitude of molecular diagnostic procedures, generic analysis that can provide several clinically relevant readouts will likely become even more important in the near future. In this context, a broad sequencing-based assay, like RNA-sequencing, to generate generic transcriptome data, from which a multitude of different readouts can be derived, presents an attractive alternative. In addition to the current study suggesting that RNA-sequencing can provide benchmarked intrinsic subtype and ROR scores, RNA-sequencing has also been reported to be able to provide reliable models for current conventional BC biomarkers ^23,48,49^ and to identify expressed somatic mutations in for example *ESR1* and *PIK3CA* ^58,59^ that may be important for future clinical management. Moreover, an upfront testing of breast cancer cases irrespective of subtype would cut lead-times for molecular assay results and may also allow future implementation of prognostic/treatment predictive signatures for clinical subgroups, like TNBC and *HER2*-amplified cases, for which there are none in clinical use today.

In summary, we demonstrate the potential of RNA-sequencing as a multipurpose assay for diagnostics and treatment decision support in early breast cancer. Based on a single analysis of fresh tissue, procured at the time of diagnosis at regional pathology departments without special sampling requirements, we demonstrate the potential to derive benchmarked equivalent estimates of current clinical markers and molecular subtypes and risk assessments, which may be extended to include mutational calls for key driver genes. Importantly, a completely open assay coupled with regionally performed sequencing, as part of routine healthcare after required validation and quality assurance, may be of strong clinical and socioeconomic value. This is demonstrated by the potential to reduce current single diagnostic marker analyses, but also the clear evidence that the gene expression-based stratification can separate otherwise seemingly homogenous clinical subgroups of breast cancer into groups with clinically relevant diverse outcomes.

## Data Availability

All data produced in the present study will be made available upon reasonable request to the authors or online upon peer-reviewed publication.

## Acknowledgements

The authors would like to acknowledge patients, clinicians, and hospital staff participating in the SCAN-B study, the staff at the central SCAN-B laboratory at Division of Oncology, Lund University, the Swedish National Quality Register for Breast Cancer (NKBC), Regional Cancer Center South, the South Swedish Breast Cancer Group (SSBCG), and the Oslo Breast Cancer Research Consortium (OSBREAC).

## Supplemental Tables 1-6

**Supplemental Table 1.**
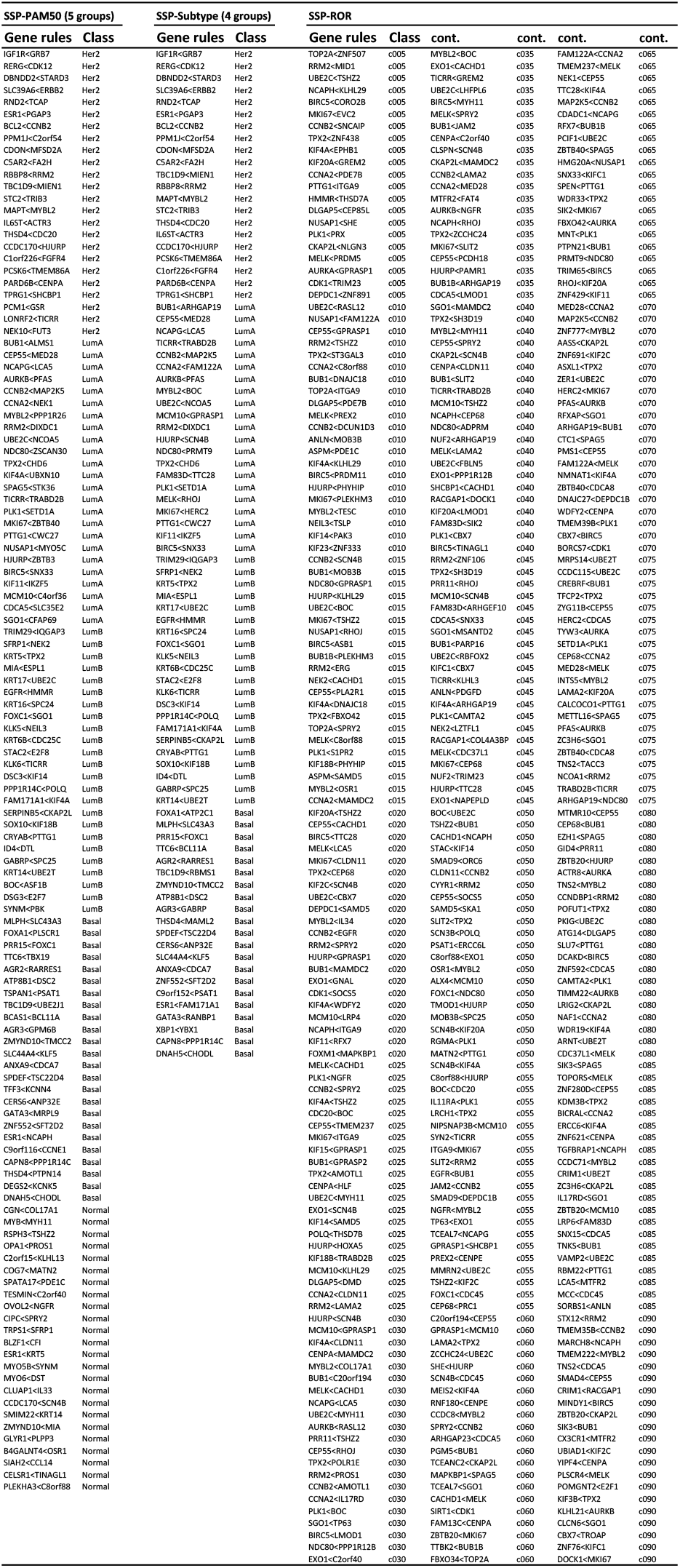
Selected SSP gene-rules for SSP-PAM50 (five groups) model, SSP-Subtype (four groups) model and SSP-ROR model.

**Supplemental Table 2.**
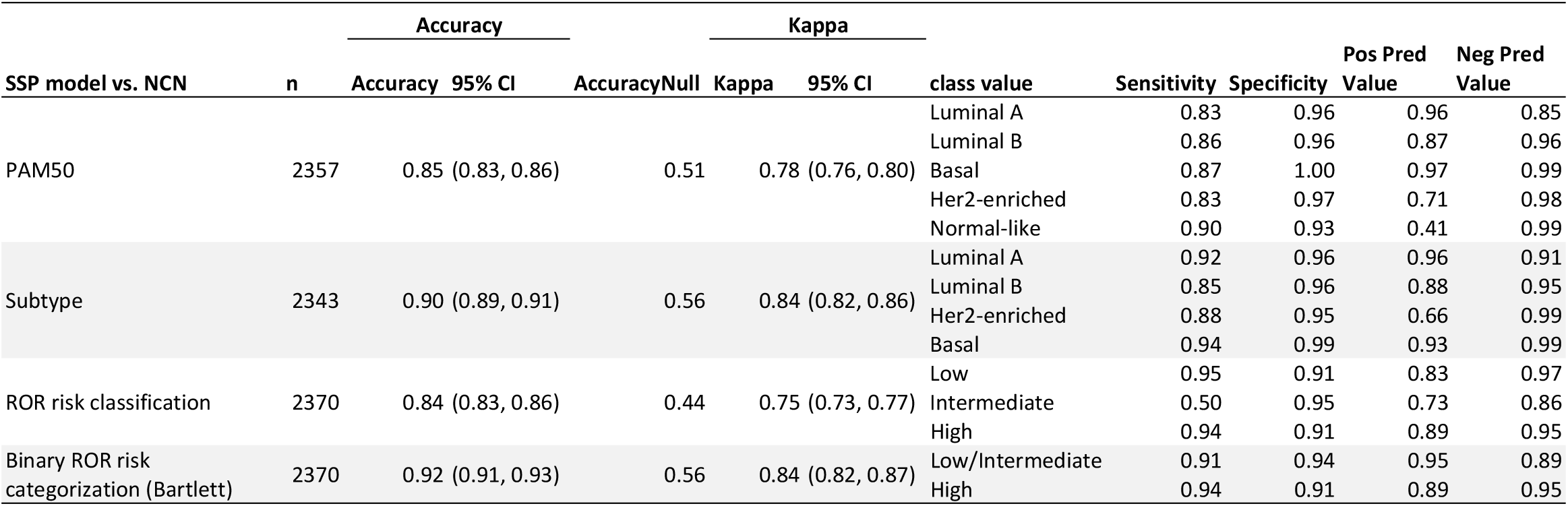
SSP prediction performance for subtypes and ROR categories validated against NCN classification in the independent population-based test set of early breast cancer.

**Supplemental Table 3.**
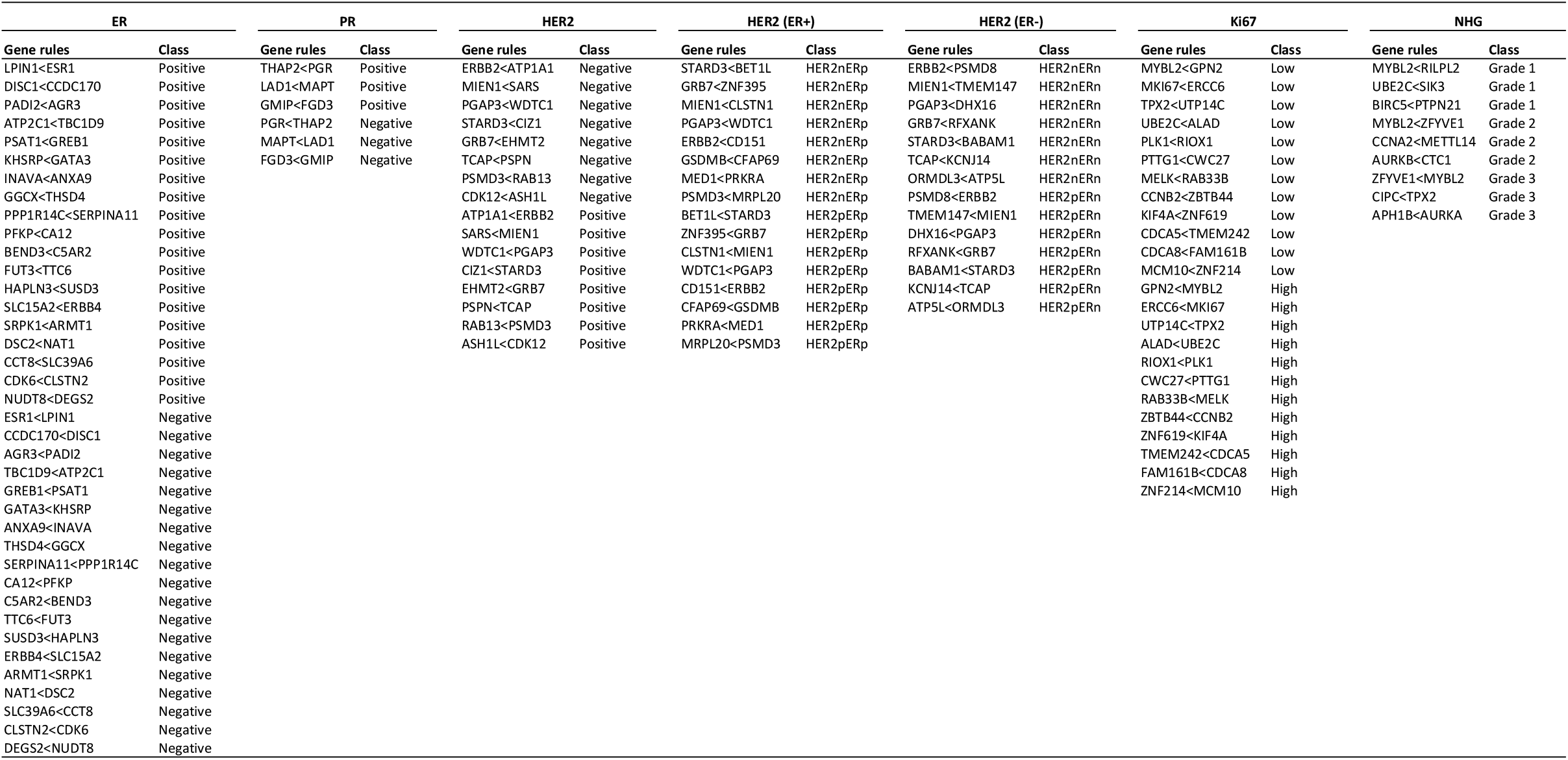
Selected SSP gene-rules for SSP models for clinical markers.

**Supplemental Table 4.**
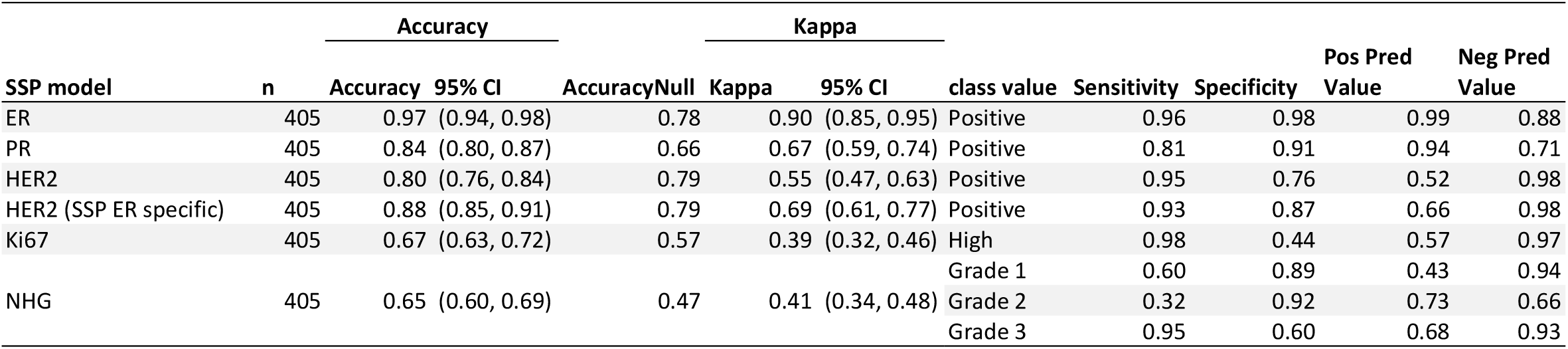
SSP performance validated against clinicopathological consensus data (re-stain and re-assessed) in the independent ABiM material from Brueffer et al. ^23^.

**Supplemental Table 5.**
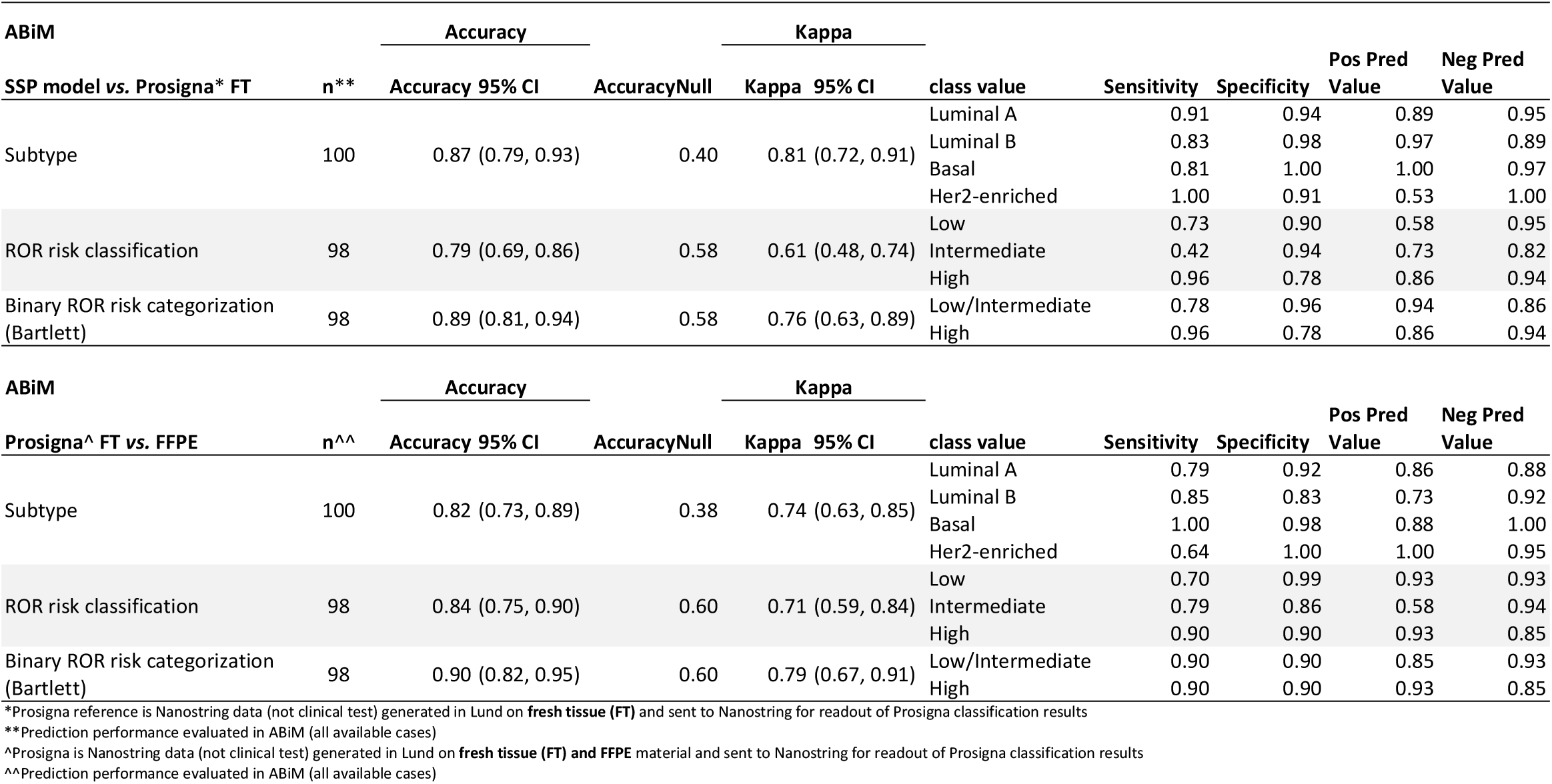
SSP model vs. Prosigna model when evaluating identically sourced RNA and Prosigna model evaluation using fresh frozen tissue (FT) vs. FFPE material in the independent external clinical series ABiM.

**Supplemental Table 6.**
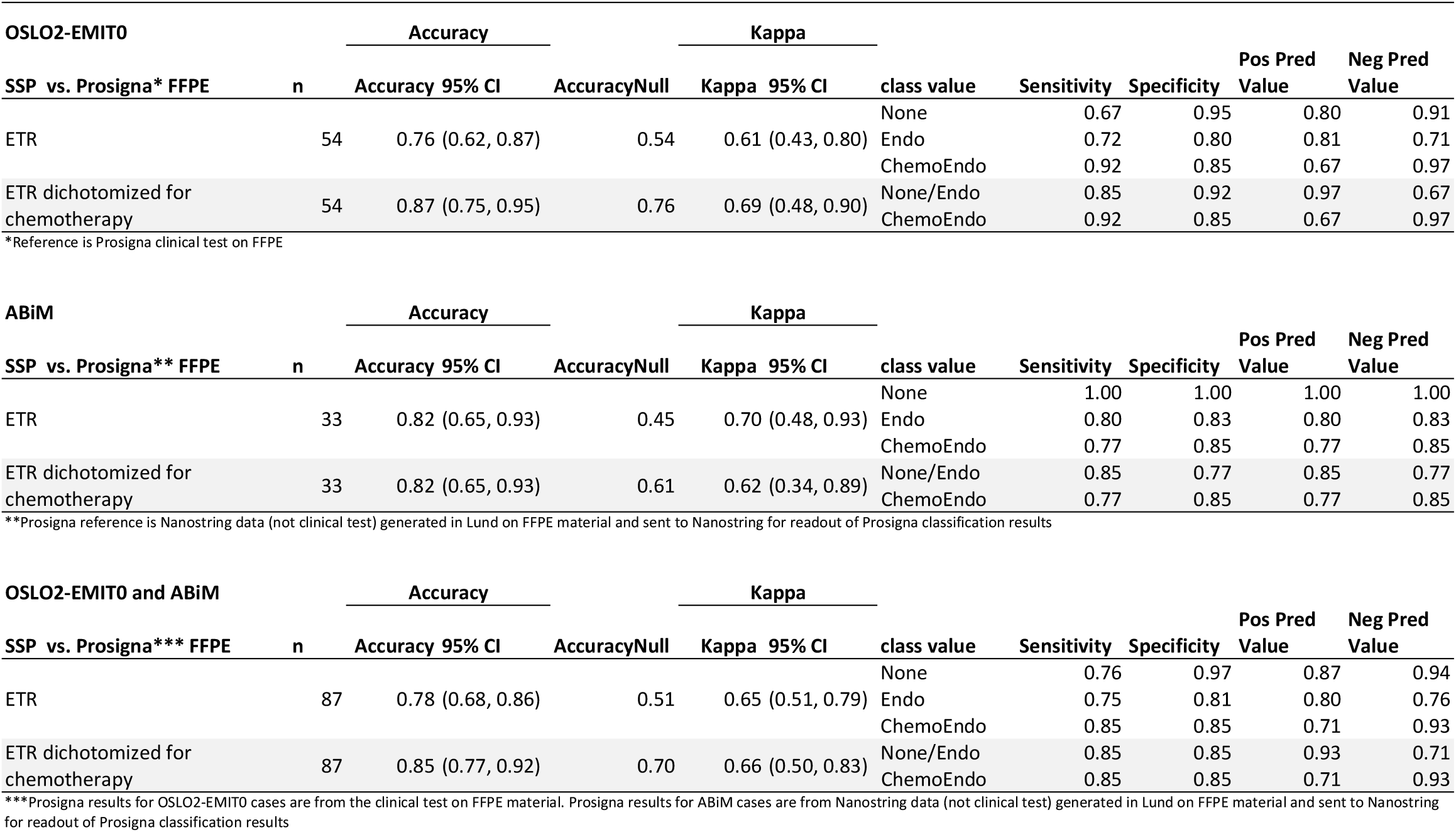
Concordance analysis of emulated treatment recommendation between SSP and Prosigna for ER+ HER2-N0 cases in the external clinical series OSLO2-EMIT0 and ABiM.

## Supplemental Figures 1-8

**Supplemental Figure 1.**
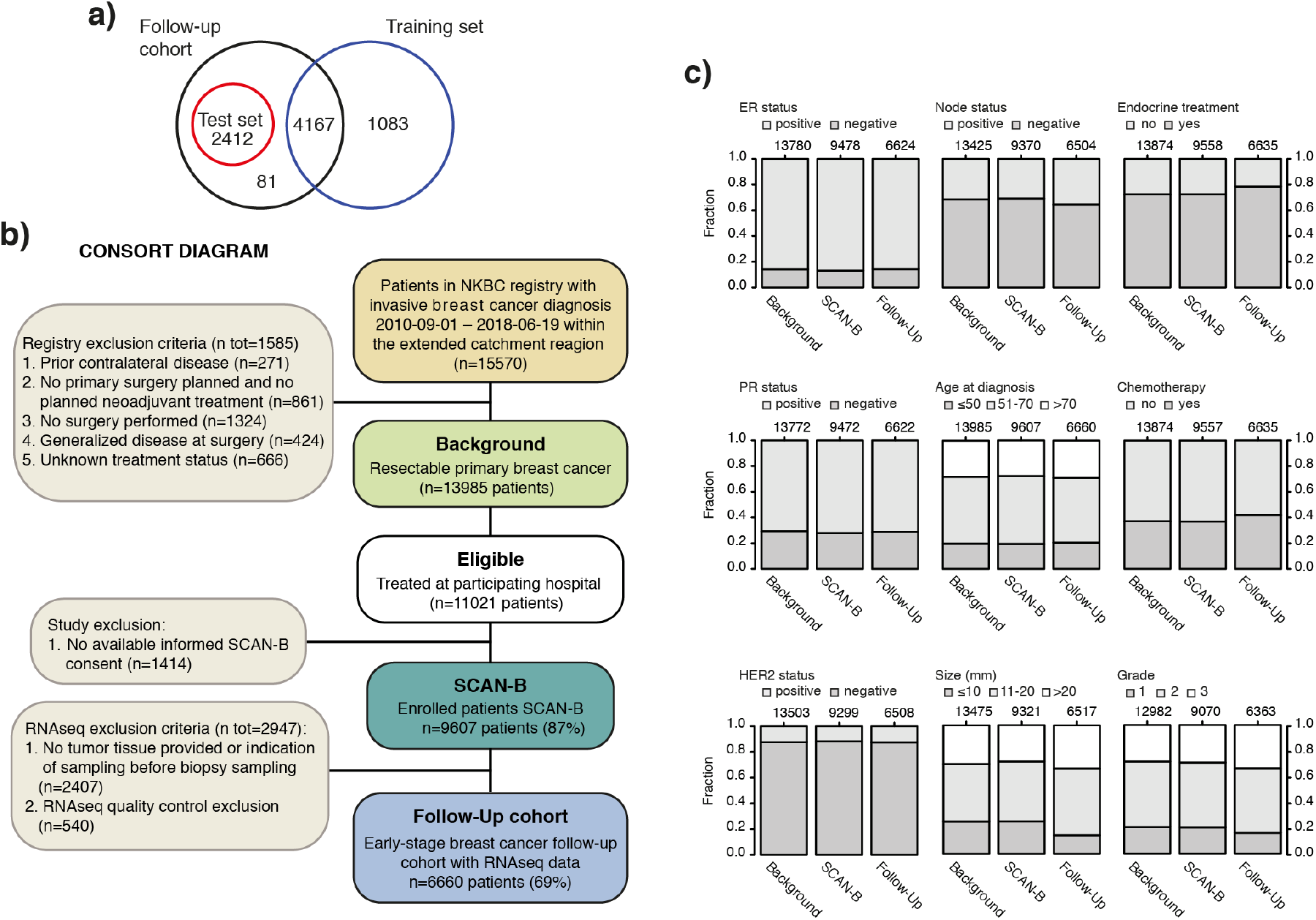
Venn diagram of patient overlap in the SCAN-B study material cohorts and consort diagram of patient selection for the early-stage follow-up cohort with bar charts illustrating population-based representativeness compared to the background population. **(a)** Venn diagram of patient overlap for the follow-up cohort of early breast cancer (black), the training set (blue), and independent test set (red). **(b)** Consort diagram of patient inclusion in the follow-up cohort of early breast cancer. **(c)** Bar charts of incidence for important clinicopathological variables in the background population (left), enrolled SCAN-B patients (center), and SCAN-B patients with RNA-sequencing data (right) in the follow-up cohort. NKBC: Swedish national breast cancer quality registry.

**Supplemental Figure 2.**
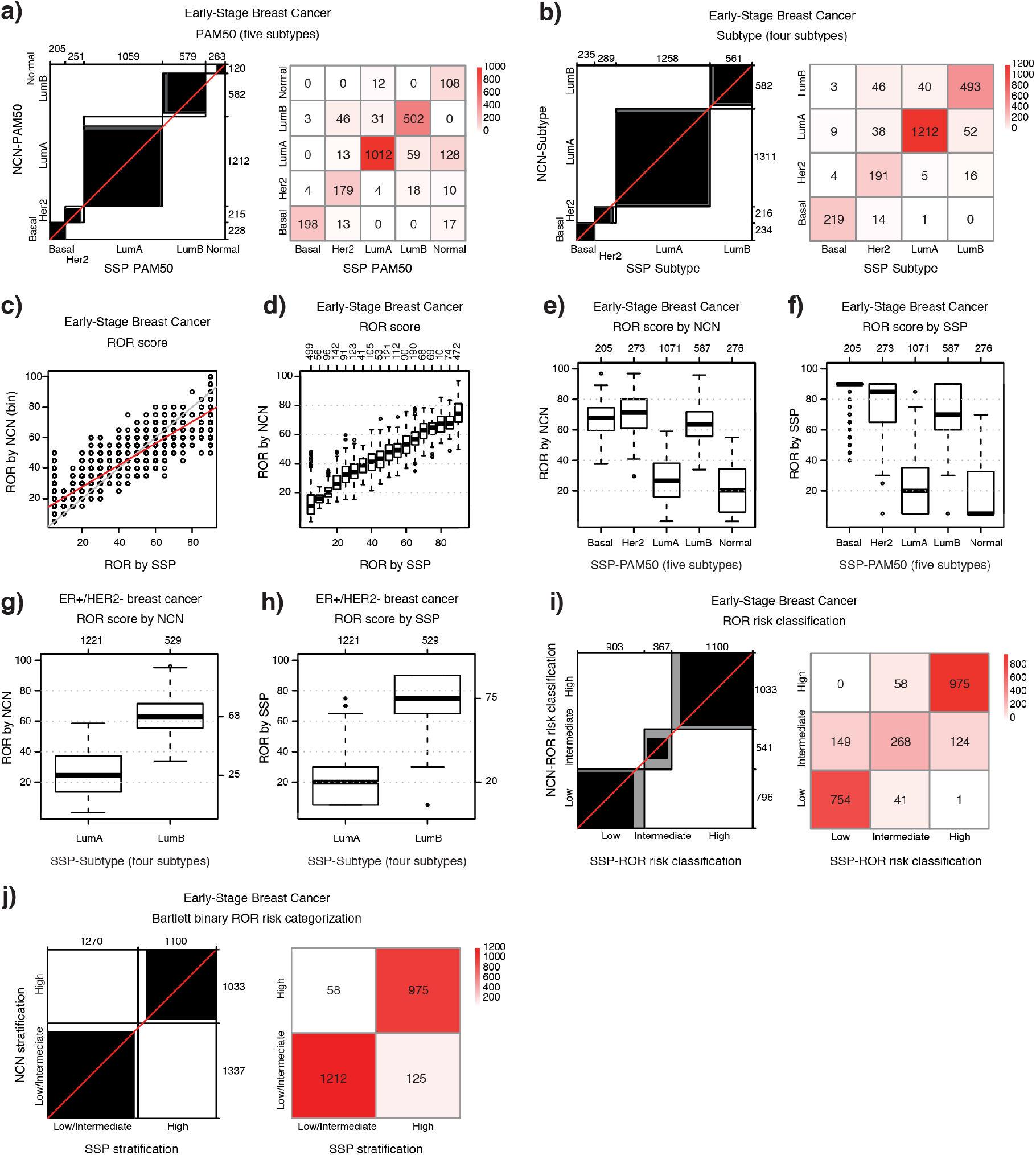
Validation of SSP classifications against NCN classifications in the independent test set of early breast cancer. **(a)** Agreement chart and confusion matrix comparing SSP classifications (x-axis/ columns) with NCN classifications (y-axis/rows) for PAM50 (five subtypes) and **(b)** Subtype (four subtypes). **(c)** Scatterplot of binned NCN-ROR values (y-axis) versus SSP-ROR (x-axis). **(d)** Boxplot of NCN-ROR values (y-axis) by SSP-ROR (x-axis). **(e)** Distributions of NCN-ROR values and **(f)** SSP-ROR values by SSP-PAM50 (five subtypes). **(g)** Distributions of NCN-ROR values or **(h)** SSP-ROR values by SSP-Subtype (four subtypes) for ER+/HER2-breast cancer classified as Luminal A or Luminal B. **(i)** Agreement chart and confusion matrix comparing SSP classification (x-axis/ columns) with NCN classification (y-axis/rows) for ROR risk classification and **(j)** emulated treatment recommendation.

**Supplemental Figure 3.**
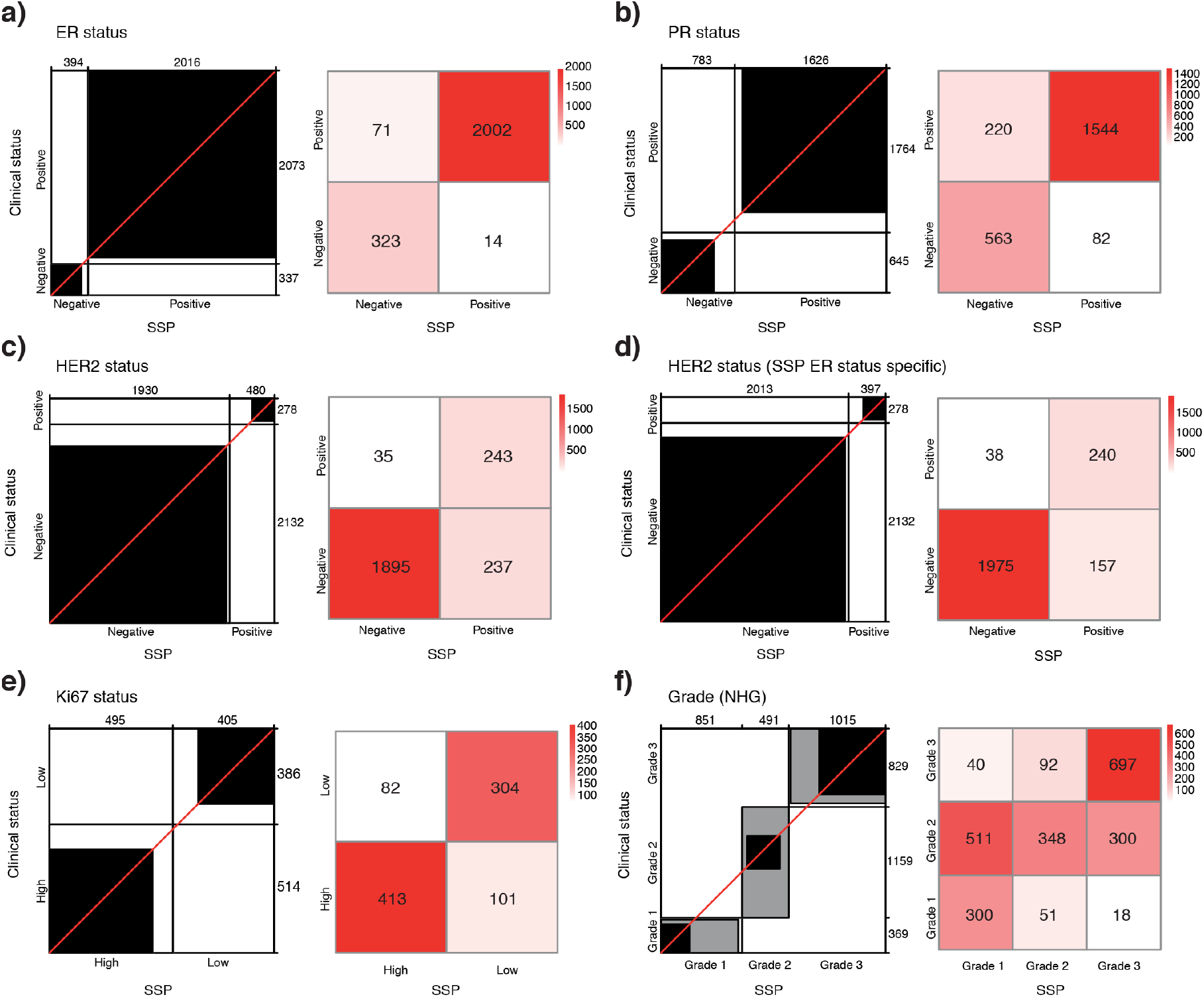
Validation of SSP models for clinical markers in the independent population-based test set of early breast cancer. Agreement chart and confusion matrix comparing the SSP classifications (x-axis/columns) with clinical histopathology status (y-axis/rows) for **(a)** ER status, **(b)** PR status, **(c)** HER2 status using a general SSP model, **(d)** HER2 status using a SSP model specific for ER status, **(e)** Ki67-status, **(f)** NHG.

**Supplemental Figure 4.**
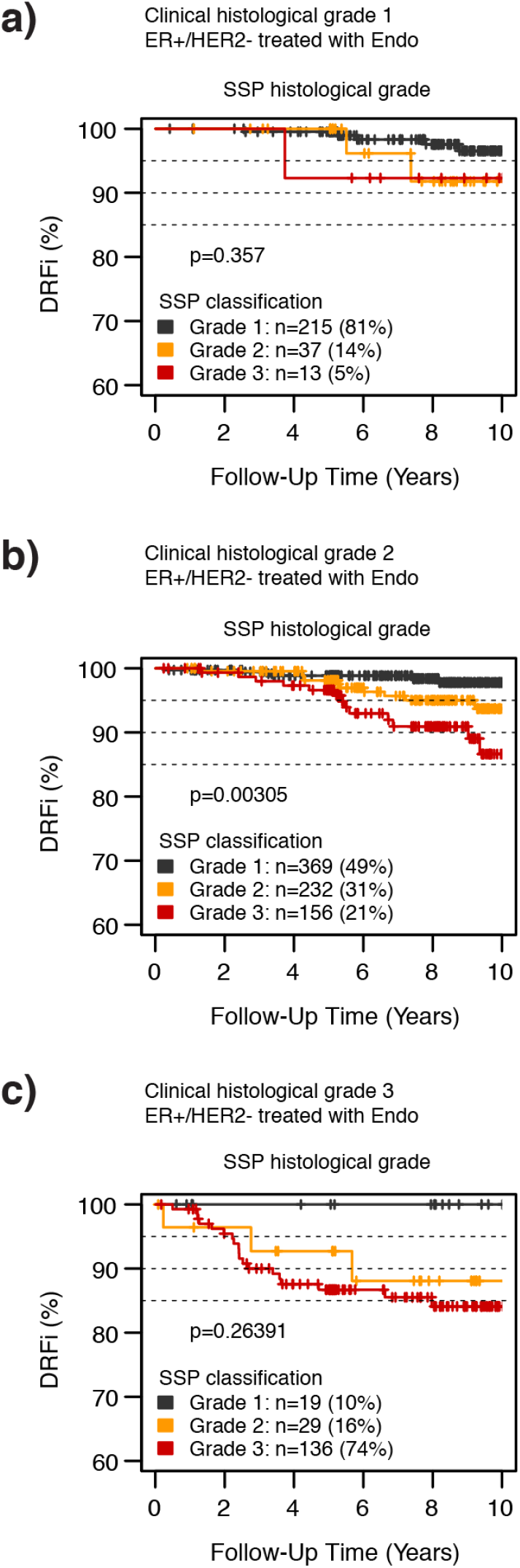
Kaplan-Maier plot for SSP classification of NHG for groups of separate clinical NHG grade in patients diagnosed with ER+/HER2-breast cancer and that only received endocrine adjuvant treatment: **(a)** clinical NHG 1, **(b)** clinical NHG 2, **(c)** clinical NHG 3.

**Supplemental Figure 5.**
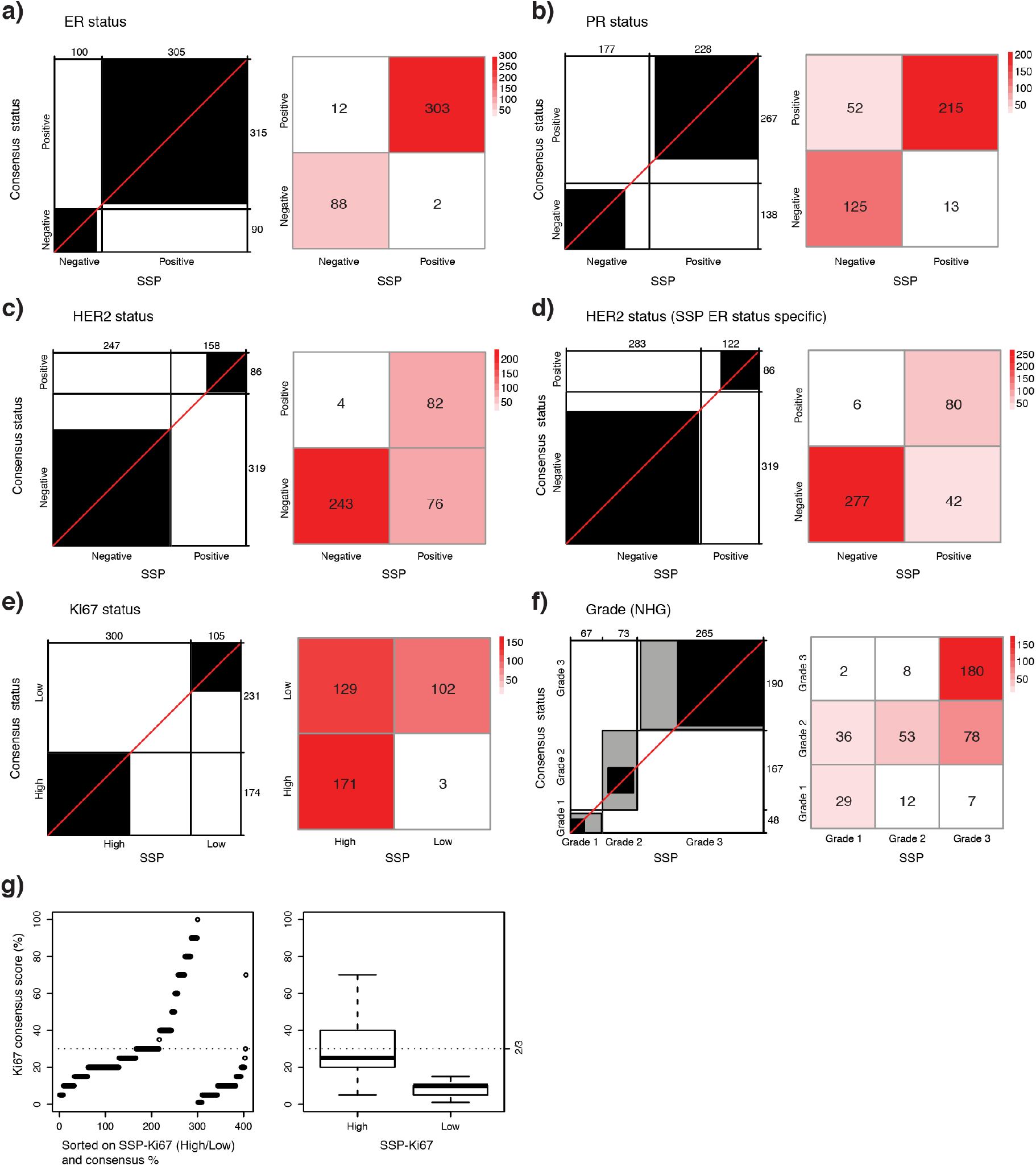
Validation of SSP models for clinical markers against consensus status in the external clinical ABiM series. Agreement chart and confusion matrix comparing the SSP classifications (x-axis/ columns) with consensus histopathology status (y-axis/ rows) for **(a)** ER status, **(b)** PR status, **(c)** HER2 status using a general SSP model, **(d)** HER2 status using a SSP model specific for ER status, **(e)** Ki67-status, **(f)** NHG, **(g)** Ki67 consensus scoring (%) by SSP Ki67 classification: samples plotted by SSP classification (left plot) and box-plots (right plot).

**Supplemental Figure 6.**
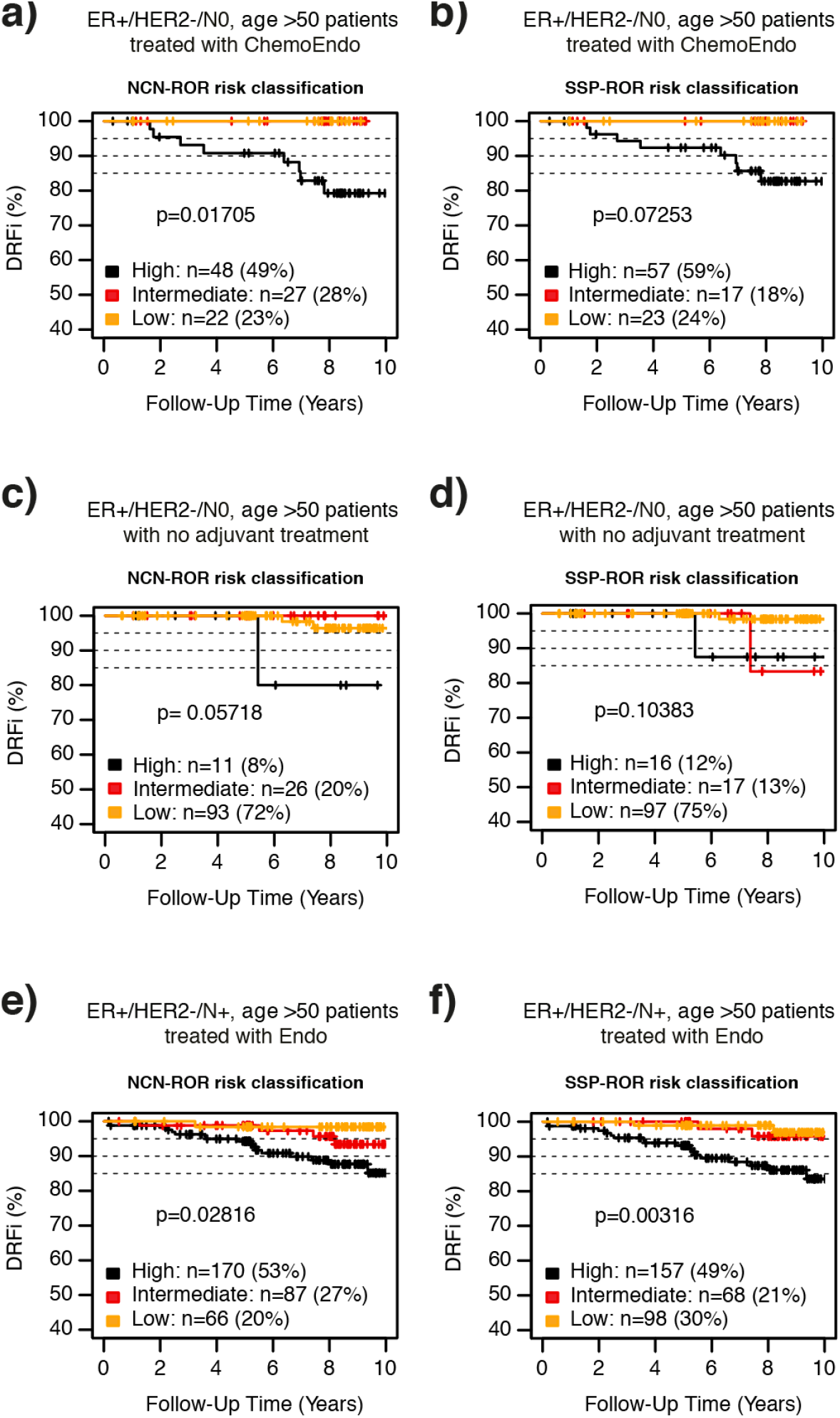
Comparing SSP and NCN classifications in the independent population-based test set by assessment of prognostic value. **(a)** Kaplan-Meier plots for stratification by ROR risk classification from SSP-ROR risk category and **(b)** NCN-ROR risk category in ER+/HER2-/N0 disease diagnosed over 50 years of age treated with chemotherapy in addition to endocrine adjuvant treatment. **(c)** Kaplan-Meier plots for stratification by ROR risk classification from SSP-ROR risk category and **(d)** NCN-ROR risk category in ER+/HER2-/N0 disease diagnosed over 50 years of age that did not receive any adjuvant treatment. **(e)** Kaplan-Meier plots for stratification by ROR risk classification from SSP-ROR risk category and **(f)** NCN-ROR risk category in ER+/HER2-/N+ disease diagnosed over 50 years of age treated with endocrine adjuvant treatment alone.

**Supplemental Figure 7.**
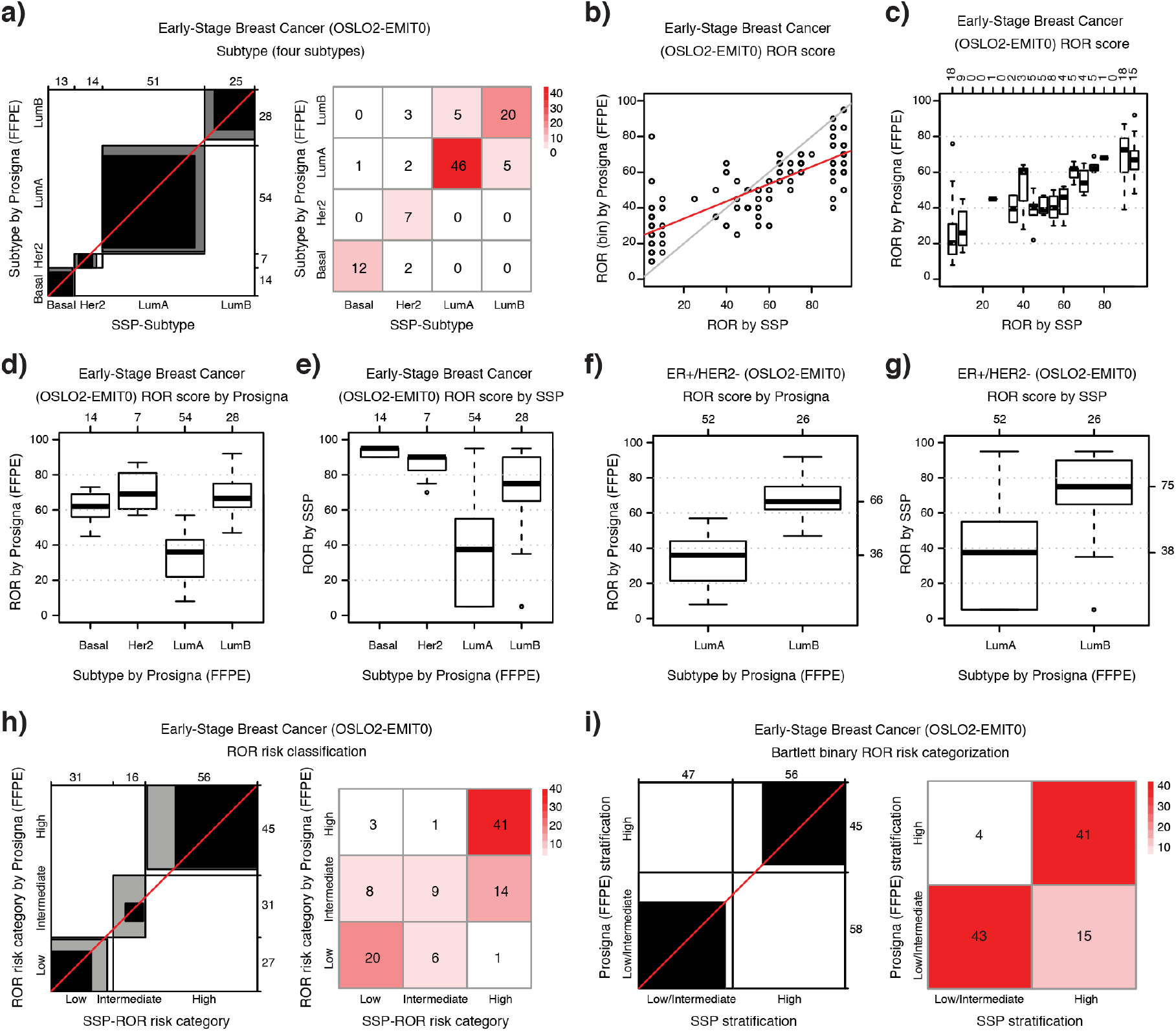
Comparing SSP classifications from fresh frozen tissue against the clinical Prosigna test from FFPE tissue in an independent clinical series of primary breast cancer (OSLO2-EMIT0). **(a)** Agreement chart and confusion matrix comparing SSP classifications (x-axis/columns) with Prosigna classifications (y-axis/rows) for Subtype (four subtypes). **(b)** Scatterplot of binned Prosigna ROR values (y-axis) versus SSP-ROR (x-axis). **(c)** Boxplots of Prosigna ROR values (y-axis) by SSP-ROR (x-axis). **(d)** Distributions of Prosigna ROR values and **(e)** SSP-ROR values by Prosigna classification for Subtype (four subtypes). **(f)** Distributions of Prosigna ROR values and **(g)** SSP-ROR values by Prosigna classification for Subtype (four subtypes) for ER+/HER2-breast cancer classified as Luminal A or Luminal B. **(h)** Agreement chart and confusion matrix comparing SSP classification (x-axis/columns) with Prosigna classification (y-axis/rows) for ROR risk category and **(i)** the two-group ROR stratification according to Bartlett et al. ^5^.

**Supplemental Figure 8.**
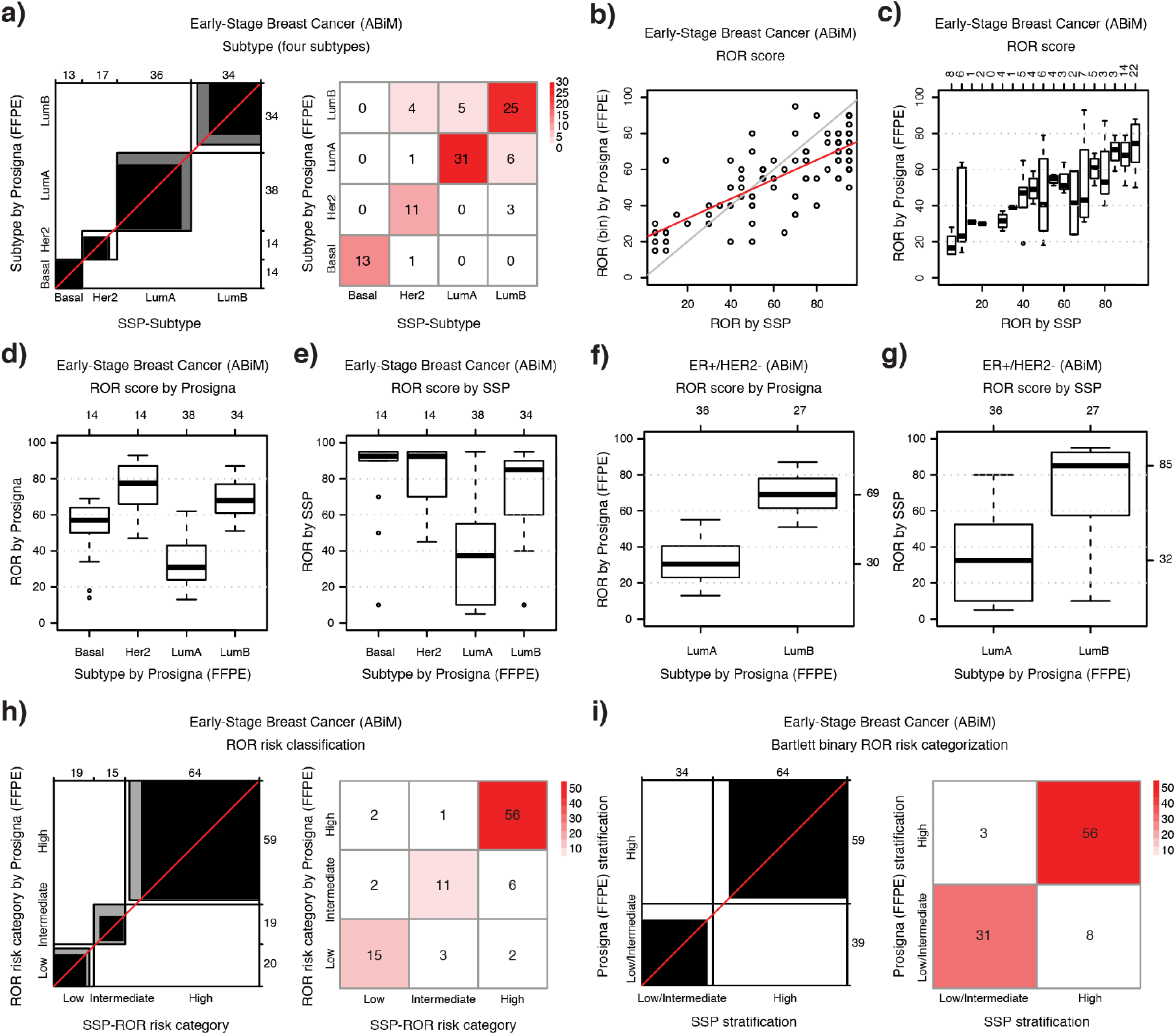
Comparing SSP classifications from fresh frozen tissue against the Prosigna model on Nanostring data from FFPE tissue in an independent clinical series of early breast cancer (ABiM). **(a)** Agreement chart and confusion matrix comparing SSP classifications (x-axis/columns) with matched Prosigna classifications (y-axis/ rows) for Subtype (four subtypes). **(b)** Scatterplot of binned Prosigna ROR values (y-axis) versus SSP-ROR (x-axis). **(c)** Boxplots of Prosigna ROR values (y-axis) by SSP-ROR (x-axis). **(d)** Distributions of Prosigna ROR values and **(e)** SSP-ROR values by Prosigna classification for Subtype (four subtypes). **(f)** Distributions of Prosigna ROR values and **(g)** SSP-ROR values by Prosigna classification for Subtype (four subtypes) for ER+/HER2-breast cancer classified as Luminal A or Luminal B. **(h)** Agreement chart and confusion matrix comparing SSP classification (x-axis/columns) with Prosigna classification (y-axis/rows) for ROR risk category and **(i)** the two-group ROR stratification according to Bartlett et al. ^5^.

