## Supplemental Methods for "RNA Sequencing-Based Single Sample Predictors of Molecular Subtype and Risk of Recurrence for Clinical Assessment of Early-Stage Breast Cancer"

#### *SCAN-B cohort*

##### *Feasibility of RNA sequencing (RNAseq) in a clinical setting with short turnaround time*

SCAN-B is an observational study with population-based inclusion in that all patients with breast cancer or suspected breast cancer within the catchment area are asked to participate. The SCAN-B material has been accumulated prospectively by continuous patient enrollment and collection of tissue since 2010. Inclusion rates are very high (>90%) over the years and the unbiased nature of inclusion is verified through comparison to the national health care quality assurance registry for breast cancer (NKBC). Whereas collected tissue has been processed with generally very short lead-times from the start of SCAN-B, extracted RNA was initially stored awaiting implementation of the SCAN-B laboratory procedures for RNAseq. When such procedures had been established and tested, the stockpiled RNA was processed and sequenced. Once the backlog of stored RNA was processed the SCAN-B laboratory transitioned to implement real-time continuous processing of collected tissue and RNAseq with short turnaround time. The real-time processing was successfully launched during the fall of 2015 and was in continuous operation until early 2019. Throughout this period, the general turn-around time from tissue arriving at the SCAN-B laboratory to the completion of RNAseq and compiling a result report with gene expression readouts was successfully maintained at less than 1 week (Supplemental methods Figure 1). Since SCAN-B activities were confined to research use only, the turn-around requirements were relaxed during summer and winter holidays, i.e., July/August and December/January (see Supplemental methods Figure 1). Nonetheless, three years of real-time processing by the SCAN-B laboratory demonstrated the feasibility in meeting turnaround

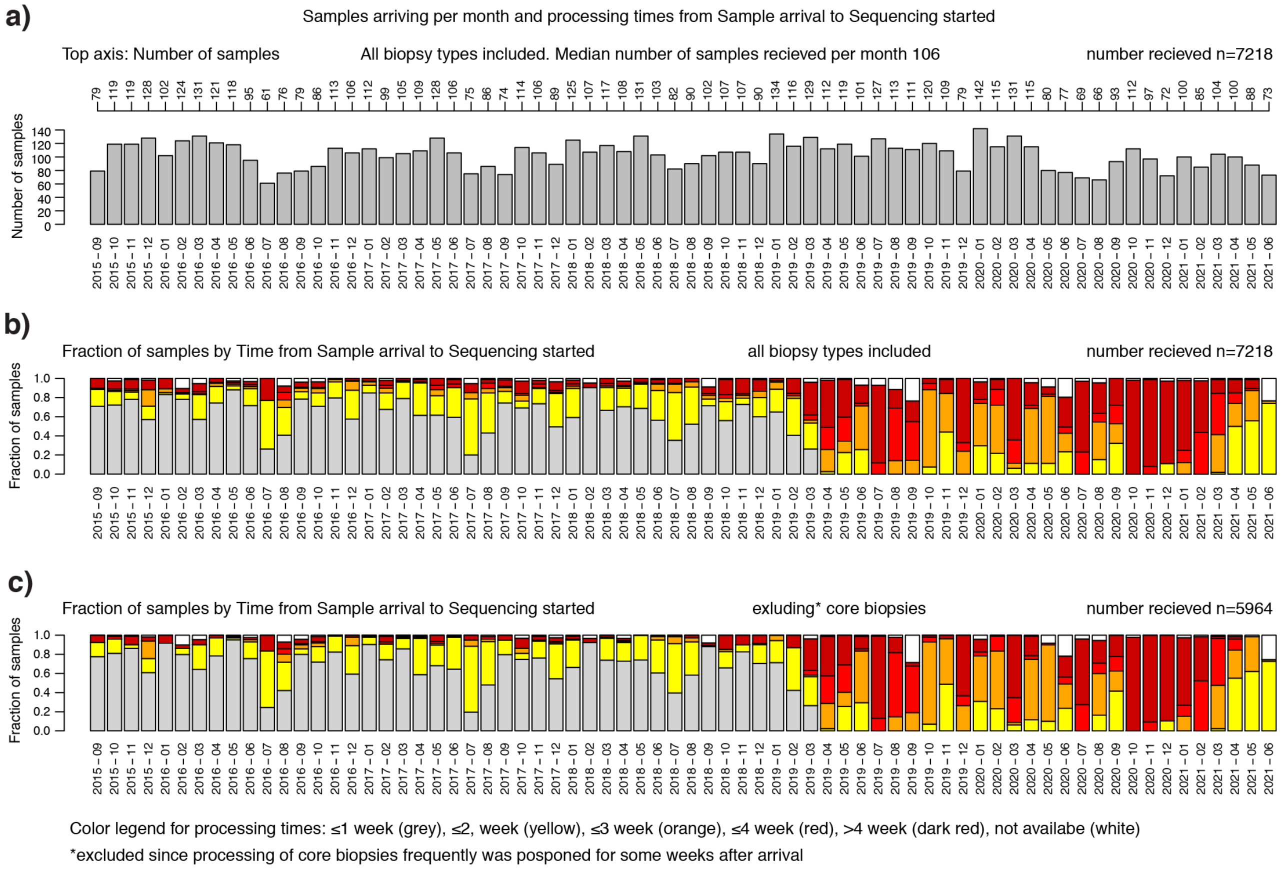

**Supplemental methods Figure 1:** Number of samples received per month at the SCAN-B laboratory and processing times (September 2015 to June 2021). Real-time processing with short turn-around times was in operation at the SCAN-B laboratory between September 2015 and early 2019. **(a)** Number of samples arriving per month including pre-operative core biopsies and biopsied taken from surgery. **(b)** Fraction of samples by processing time from arrival to sequencing including both core biopsies and biopsies taken from surgery. **(c)** Fraction of samples by processing time from arrival to sequencing for biopsies taken from surgery specimen (excluding core biopsies). In general, the time required from sequencing started to the completion of automated demultiplexing and creation of compiled GEX results reports was <24h. Color coding for processing time: ≤1 week (grey), ≤2, week (yellow), ≤3 week (orange), ≤4 week (red), > 4 week (dark red), not available (white).

time requirements for clinical use regarding tissue sampling and RNAseq. From 2019 the SCAN-B laboratory has continued processing breasts cancer tissue samples but without adhering to short lead-times.

*Study-specific cohort stratification*

The SCAN-B material used in this study includes a total of 9206 RNAseq derived gene expression profiles (GEXs). The material was divided into separate study cohorts used for training and test/validation. A test cohort suitable for follow-up analysis was selected. The test cohort comprised 2412 patients diagnosed with early-stage surgically resected breast cancer between September 1 2010 and December 31 2013 and with available clinicopathological data for stratification into relevant clinical evaluation groups based on classic clinical markers and treatment. There are no technical replicates included in the test cohort and it is non-redundant, i.e., each included patient is represented by a single GEX. Second, an isolated training cohort was set apart comprising 5857 GEXs from 5711 tissue samples derived from 5250 patients. Most importantly, all training of SSP models was completely confined to the training set and there is no patient overlap between the training cohort and the test cohort. In that respect, the test set is completely independent from the training material and validation can be regarded as independent.

#### *Gene expression data*

Gene expression data was derived from RNAseq data from Illumina sequencers using a SCAN-B analysis pipeline based on open source software. Analysis steps were implemented as an automated analysis pipeline in BASE [1] with extension package Reggie (BioRxiv: https://doi.org/10.1101/038976). GRCh38 [2] [grch38], dbSNP [3] [dbsnp], and GENCODE [4] [gencode] are used to create alignment and transcript targets. The analysis steps are listed in sequence (default software parameter values are used except as stated below):

1. Raw base call data is generated with Illumina HiSeq, NextSeq, and NovaSeq sequencers according to manufacturer's instructions (Illumina).
2. Demultiplexing the raw sequencing reads is done with Picard tools [5] ExtractIlluminaBarcodes and IlluminaBasecallsToFastq. Several versions of Picard tools have been used 1.120, 1.128, 2.20.0, 2.20.8, and 2.22.3. Only PF annotated reads are retained during demultiplexing.
3. The demultiplexed sequence files are processed with trimmomatic [6] to remove parts of the readouts that match adapter sequences and to remove poor quality base pair readouts at the end of the reads. Two versions of trimmomatic have been used, versions 0.32 and 0.33. Trimmomatic is run in two passes. First we filter adapters with parameters `ILLUMINACLIP:TruSeq3-PE-2.fa:2:30:12:1:true` and `MINLEN:20`. The second pass is then run with parameters `MAXINFO:40:0.9` and `MINLEN:20`.
4. Reads that match a target of selected genomic sequences are removed. This filtering step remove reads that are considered to be of no use in subsequent analysis – spiked in Enterobacteria phage phiX174 (RefSeq identifier NC_001422.1), ribosomal RNA (RefSeq NR_023363.1, NR_003285.2, NR_003286.2, NR_003287.2, and Genbank X12811.1, U13369.1), repeating sequencing elements (RepeatMasker@UCSC Genome Browser hg38 [2] [genomebrowser]). Bowtie [7] was used as the aligner in this filter step. Several 2.2.x versions of bowtie have been used. Key parameters used are: `--fr -k 1 --local`
5. Alignment is performed by Hisat version 2.1.0 [8]. The GRCh38 human genome assembly is used as the genome target and the transcriptome target is defined by the GENCODE release 27 [4]. The hisat index is augmented using the `--snp` parameter during index creation. dbSNP build 150 is used for all alignments. Key parameters used are: `--fr --no-unal --non-deterministic --novel-splicesite-outfile aligned/splicesites.tsv --rna-strandness RF`
6. Expression estimation is performed with stringtie [9] version 1.3.3b using protein coding transcripts from GENCODE release 27 as transcriptome model. Novel transcripts are discarded. Key parameters used are: `--rf -e`
7. Summary (merge on gene symbol). For collapsing on gene, the sum of expression values of each contributing transcript is calculated.

#### *Assigning PAM50 subtype and ROR score using nearest centroid*

*Nearest centroid (NC) classification and NCN classification.*
PAM50 subtype and ROR score were assigned by nearest centroid (NC) classification using published PAM50 subtype centroids, and largely following the general strategy previously established by Parker et al. [10], i.e., using a reference set of samples for appropriate gene transformation before calculating correlations to PAM50 centroids.

In order to correctly transform gene expression to match the training population from which the PAM50 centroids were originally derived we selected the reference samples by matching the clinicopathological metadata of samples in the original training population defined by Parker et al. to SCAN-B cases. In addition, we extended the general strategy by utilizing multiple reference sets of samples to achieve an NCN classification, i.e., nearest centroid with N reference sets and with N=100. Thus, the reference set selection procedure was repeated 100 times to create a series of 100 static reference sets, each matched to mimic the original training population.

*Candidates for selecting NCN reference sets*
A subset of the training cohort was chosen as candidates for NCN reference set selection. The candidate cohort was selected to be non-redundant for case (i.e., patient/laterality) and to only include GEX data from samples from invasive breast cancer with available clinicopathological metadata relevant for matching the original PAM50 training population from Parker et al. Included GEX data was also required to meet quality control criteria defined below.

Available clinicopathological metadata requirements:

- Invasive breast cancer
- ER status
- HER2 status
- Lymph node status
- Nottingham grade (NHG) status

RNA and GEX quality control criteria:

- RNA QC (abs) ≥6
- (ALIGNED ≥10M AND fraction duplicates ≤ 0.55) OR UNIQUE READS ≥10M

When multiple GEX assays where available for a case, one assay was randomly selected from the RNA with the highest original concentration by the rational that higher RNA yield would indicate higher tumor cellularity. The resulting cohort of candidates (pam50.reference.pre.cohort) included GEX for a total of 3334 samples distributed between different library protocols as follows: dUTP n=1107, NeoPrep n=938, and TruSeq manual n=1289.

*Library protocol correction*
GEX data from different library protocols is expected to affect relative gene expression. As centroid classification is relativistic, we constructed a simplistic library protocol adjustment procedure to transform GEX from one protocol-base to another. We first stratified data by library protocol while controlling for clinical subgroups so that clinical subgroup proportions were identical across each stratum. We reasoned that the average gene expression for individual genes should be the same in groups balanced for clinical subgroups when selected from the same overall clinical series. We calculated average gene expression for each gene in respective collection and used the gene specific delta (e.g., see truseq.dutp.diff below) between strata to adjust gene expression from one library protocol to another. Thereby, all gene expression data could be transformed to TruSeq-like expression as a common baseline. We performed SWAMP [11] analysis before and after library protocol correction and confirmed that library protocol signal in the gene expression data was removed, while signal to relevant clinical variables were retained (Supplemental methods Figure 2).

To construct the library protocol adjustment procedure, we used the candidates for NCN reference set selection, i.e., the pam50.reference.pre.cohort. From this data we created three separate cohorts, each with two groups of library protocols (dUTP/TruSeq, dUTP/NeoPrep, and NeoPrep/TruSeq, respectively) to derive gene specific mean differences between library protocols (Supplemental methods Table 1). Again, the assumption was that by having balanced groups any difference in gene

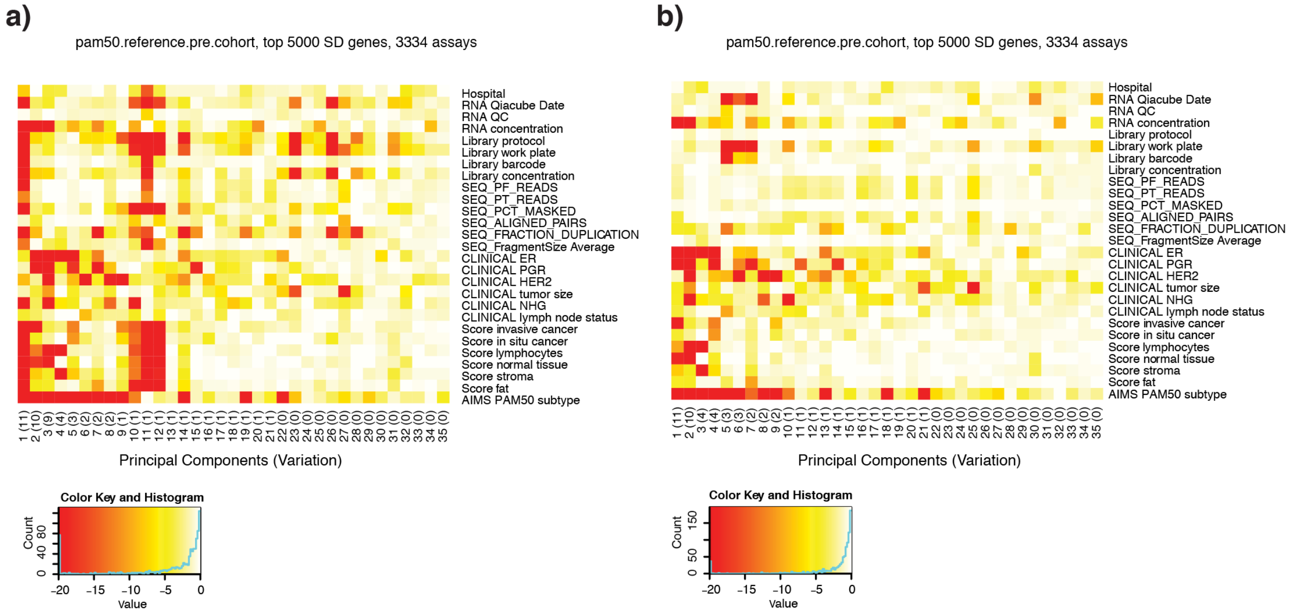

**Supplemental methods Figure 2.** SWAMP analysis before and after library protocol correction. Analysis is done using the 5000 genes with largest standard deviation measured across the analyzed cohort. **(a)** Analysis before library protocol correction. **(b)** Analysis after library protocol correction.

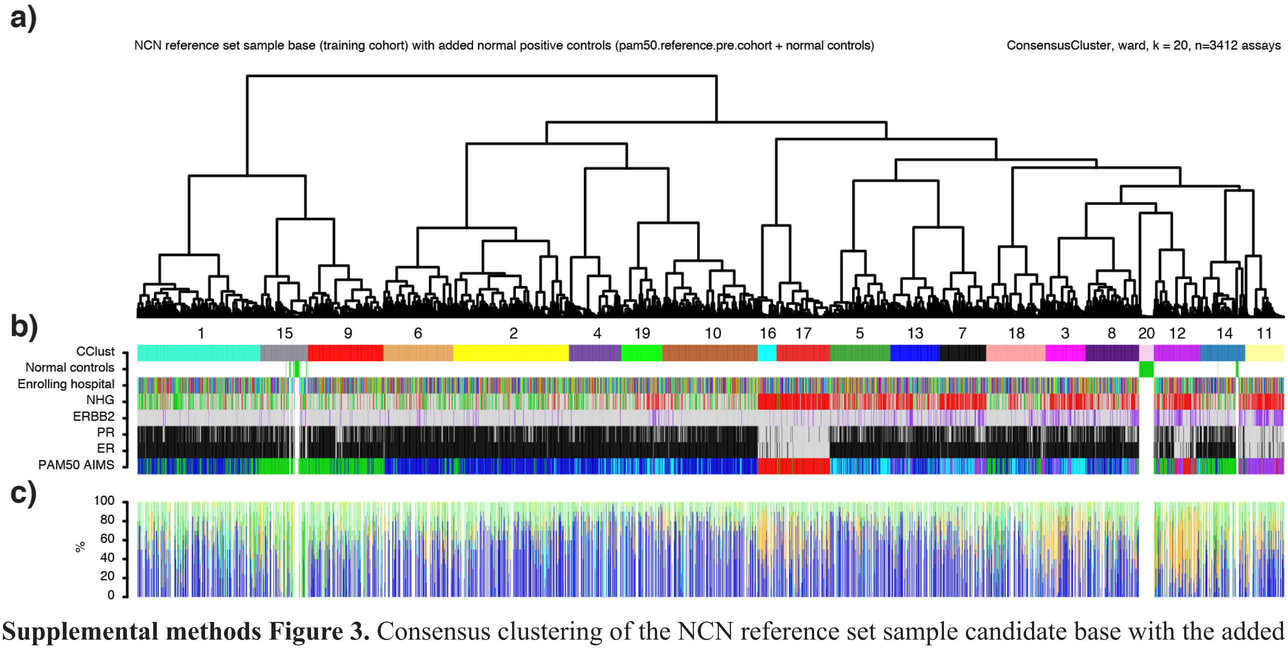

**Supplemental methods Figure 3.** Consensus clustering of the NCN reference set sample candidate base with the added normal positive GEX controls (pam50.reference.pre.cohort + normal controls) using transposed eigenvectors from PCA analysis of gene expression of all genes (>19000). **(a)** Dendrogram from consensus clustering with k=20. Consensus cluster number (1-20) is shown below the dendrogram. **(b)** GEX assay annotations. CClust: Consensus cluster. Normal controls: normal tissue control (green). Enrolling hospital: SCAN-B enrollment site. NHG: Grade 1 (green), Grade 2 (grey) Grade 3 (red), not available (white). ERBB2: Positive (purple), Negative (grey), not available (white). PR: Positive (grey), Negative (black), not available (white). ER: Positive (grey), Negative (black), not available (white). AIMS: Subtypes by model from Paquet and Hallett 16, Luminal A (blue), Luminal B (cyan), HER2-enriched (purple), Basal-like (red), Normal-like (green), not available (white). **(c)** Bar plot with percentage estimate for cell type from cellularity scoring by a breast cancer pathologist: invasive (dark blue), in-situ (light blue), fat (yellow), immune cells (orange), normal mammary epithelial (dark green), stromal (light green), not available (white). Bars correspond to samples. The scored tissue was taken adjacent to the piece used for RNA-sequencing, as part of SCAN-B laboratory processing, and then formalin-fixed and paraffin-embedded to produce a low-density tissue microarray used for cellularity estimation [17].

expression mean can be attributed to the library protocol. The calculated protocol differences per gene were used as correction factor for converting gene expression between protocols.

For example, protocol difference (per gene) for TruSeq and dUTP, i.e., truseq.dutp.diff, was calculated per gene using:

```
truseq.dutp.diff = mean(log2( TruSeq+0.1 )) - mean(log2( dUTP+0.1 ))
```

The idea is that dUTP can be transformed to TruSeq using:

```
log2(TruSeq+0.1) = log2(dUTP+0.1) + truseq.dutp.diff
```

The transformation function to transform dUTP GEX to TruSeq(like) GEX per gene:

```
T = dUTP*2^truseq.dutp.diff + 0.1*(2^truseq.dutp.diff - 1)
```

To avoid negative values, transformed values were capped at 0, i.e., if $T<0$ then T was set to 0.

Similar protocol differences (per gene) were also calculated to be able to transform gene expression between TruSeq and NeoPrep and between dUTP and NeoPrep library protocols.

*Normal breast tissue controls*

Normal breast tissue samples from two sources were included and added to the candidates for NCN reference set selection. Source one included 12 GEX from normal breast tissue specimens collected by pathologists from the extended surgical resection specimen of SCAN-B breast cancer patients. Source two included 66 GEX from non-SCAN-B normal breast tissue specimens obtained from mammoplasty surgery from healthy women.

*Curating the cohort for NCN reference selection*

In order to evaluate the normal tissue samples and to further critically curate all GEX data before selecting NCN reference sets we performed a consensus clustering using transposed eigenvectors from PCA analysis of GEX including all available genes (Supplemental methods Figure 3). Consensus clustering was performed using Bioconductor package ConsensusClusterPlus [12] evaluating 20 clusters with (1 - Pearson correlation) as distance function and using ward.D2 as hierarchical linkage methods. Subsampling was repeated 800 times sampling 80% of samples (GEX) and 98% of features (genes). A number of the external mammoplasty tissue grouped by themselves in consensus cluster (CC) 20 and were excluded as outliers. A small fraction of the remaining mammoplasty tissue and of the normal tissue from SCAN-B grouped in CC 14 together with other samples of mixed clinicopathological characteristics. The remaining normal samples, of both mammoplasty and SCAN-B origin, were confined to a sub-branch of CC 15 were they intermingled with a number of predominantly ER positive, HER2 negative low-grade SCAN-B samples. All normal samples in consensus cluster 14 and 15 were considered suitable positive normal breast tissue controls, whereas other GEX that grouped in these clusters were deemed as possibly normal-contaminated and excluded from NCN reference set selection. A final curated quality controlled sample base of 3094 tumor tissue and 27 normal controls remained for NCN reference set selections. Data from the curated sample base were used to select 100 static reference sets by matching and bootstrapping. None of the reference sets overlap by more than 30%.

#### *Training SSP models using the AIMS procedure*

*Normal and SSP model for suspected low invasive cancer cellularity.*

Based on the consensus clustering results we reasoned that GEX from tumor samples that grouped in CC 15 were at especially high-risk of being affected by low cancer cellularity in the obtained tissue sample. Thus, it would therefore be beneficial, or at least not detrimental, to exclude those from SSP training. We therefore first trained an SSP model for CC 15 using *Norm15* and *Rest* as class-labels, i.e., samples that grouped in CC 15 (*Norm15*) versus samples that did not group in that cluster (*Rest*). For training the SSP-CC15 model, a total of 3346 GEX were used (Norm15 n=125 and Rest n=3221). The maximum overall agreement was observed at 27 gene-rules per group by AIMS, although only marginal improvements was seen using >2 rules. The union of unique Entrez ID in selected rules was 54. The overall accuracy in the training set was 96.9%.

To evaluate the SSP-CC15 model in an independent test set we classified a TCGA breast cancer cohort, comprising 994 breast tumors and 106 normal breast tissues analyzed by RNAseq. The SSP-CC15 successfully classified 103 of 106 normal and 980 of 994 tumors for an overall accuracy of 98.5%. We therefore considered the SSP-CC15 an appropriate tool for identifying GEX data potentially affected by low tumor cellularity that could be omitted from further SSP training.
The entire training cohort was classified using the SSP-CC15 with 452 of 5857 (8.4%) GEX classified as *Norm15*, equivalent to data from 446 of 5711 (7.8%) individual tissue samples. Correspondingly, the remaining 5405 GEX were available for further SSP training.

*Assigning NCN class-labels for intrinsic subtypes to use for SSP training*

The non-TruSeq gene expression data for the NCN static reference sets was transformed to TruSeq-like expression using the gene specific deltas (see section *Library protocol correction* of Supplemental methods). Consequently, before applying the NCN classifier, any non-TruSeq raw gene expression data from individual test samples was first adjusted to TruSeq-like for a common baseline.

Next, when applying the NCN classifier, a test sample is first normalized to a static reference set and then compared to subtype centroids by correlation. The normalization was done per gene by centering, using the mean from the reference set, after first adding 0.1 (to avoid logarithms of 0) to FPKM followed by log2 transformation. Correlation to subtype centroids was done by spearman correlation. Subtype was assigned by nearest centroid (highest correlation), requiring a minimum correlation of 0.2 or otherwise assigned as unclassified. ROR was calculated within the normalized reference set data using centroid correlations, tumor size and proliferation score by the ROR equation (below) as described [13-15], followed by scaling the range to 0-100.

$$ROR_{non\text{-}scaled} = 54.7690*(-0.0067*B + 0.4317*H - 0.3172*LA + 0.4894*LB + 0.1133*T + 0.1981*P + 0.8826)$$

B: correlation to the basal-like centroid
H: correlation to the HER2-enriched centroid
LA: correlation to the Luminal A centroid
LB: correlation to the Luminal B centroid
T: pathologic tumor size coded as 0 (≤20 mm) or 1 (>20 mm)

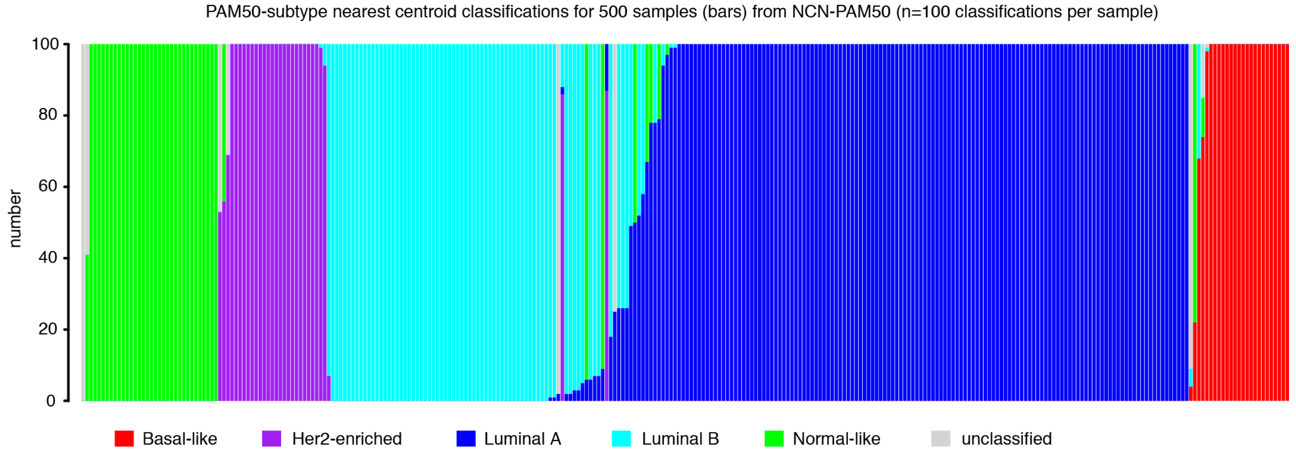

**Supplemental methods Figure 4.** Individual NCN (n=100) PAM50-subtype classifications for a random sample of 500 GEX assays from the training cohort. Bars correspond to samples.

P: proliferation score (mean expression of an 18-gene proliferation subset from Walden et al. [15] and part of the PAM50 genes)

For each test sample the process is repeated 100 times, using each static reference set for normalization. Scaling of $ROR_{non\text{-}scaled}$ value was done for respective static reference set together with the test sample. Thus, the NCN is essentially 100 separate implementations of a PAM50 classification and accordingly the output for a test sample is a series of 100 individual subtype assignments and scaled ROR scores. For assigning one NCN subtype to a test sample we used the majority vote from the 100 individual classifications and for assigning one NCN ROR score we used the average value of the 100 individual ROR scores. For final SSP training, samples assigned as unclassified by NCN majority vote were excluded.

We applied NCN-PAM50 classification to the entire training cohort to obtain class-labels for training SSP models. We created two sets of class-labels for intrinsic subtype. First, *PAM50-subtype* corresponding to the original intrinsic subtype schema. Second, *Subtype* corresponding to the Prosigna assay with only four categories, omitting the Normal-like centroid when assigning subtype. In most instances, the NCN majority vote assigned subtype with 100% consensus. However, for a number of GEX the vote was not unanimous, but a subtype could still be assigned using a clear majority, and in a small number of instances the vote was close to a draw as exemplified in Supplemental methods Figure 4. These discrepancies demonstrate the relativistic nature of nearest centroid assignments and the risk of classification variance when relying on a single static reference set.

*Assigning NCN class-labels for ROR categories to use for SSP training*

The ROR score can be related to risk of recurrence using reference patient materials with known clinical outcome. In this manner, predefined cutoffs can be derived for classification into Low, Intermediate, and High risk of recurrence. Different cutoffs are used depending on the nodal status of patients. In addition, the ROR function include the parameter gross tumor size (T) with categorical value 0 or 1 depending on the tumor size from pathologist examination: T=0 for tumors ≤20mm and T=1 for tumors >20mm. Taken together this makes it challenging to construct a categorical SSP model for ROR. To this end, we assigned categorical ROR values to samples through data binning to obtain ROR class-labels that could be used for training SSP models. To determine a suitable and practically useful bin width we related the effect of gross tumor size to the standard deviation (SD) of ROR from our NCN classification. In the training set, respective average SD ($SD_{avg}$) for ROR, i.e., average of the individual sample SD from the n=100 ROR from NCN, in the two subgroups of gross tumor size was 2.46 for T0 and 2.16 for T1 (Supplemental methods Figure 5 a). We empirically determined the effect of tumor size by calculating ROR twice using alternate settings for parameter T (0 or 1). The $SD_{avg}$ across the cohort for respective T setting was 2.39 for T=0 and 2.29 for T=1 (Supplemental methods

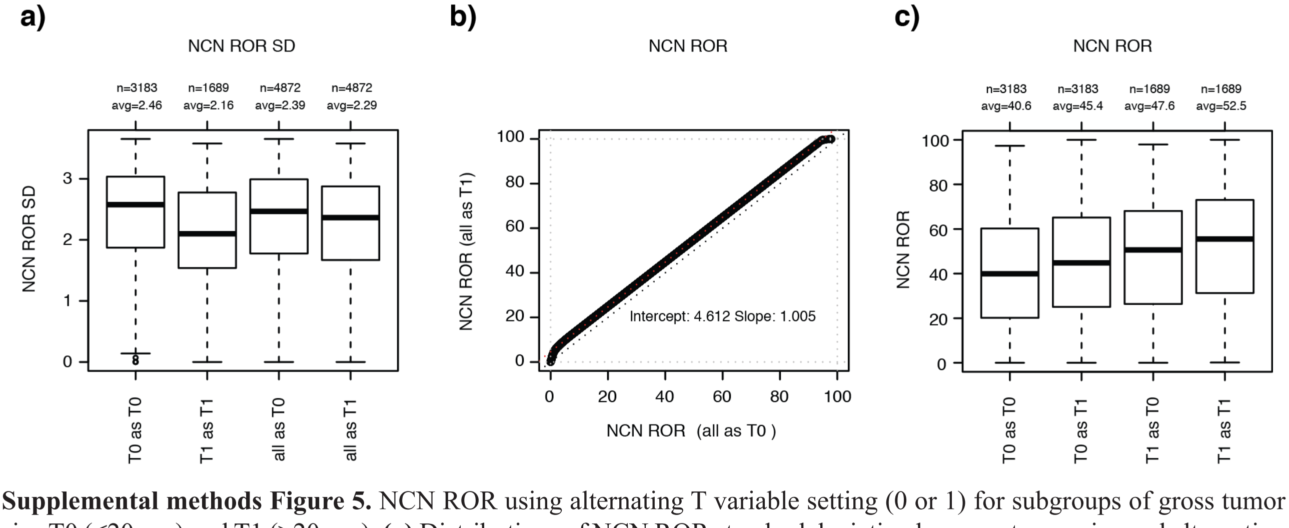

**Supplemental methods Figure 5.** NCN ROR using alternating T variable setting (0 or 1) for subgroups of gross tumor size T0 (≤20mm) and T1 (>20mm). **(a)** Distributions of NCN ROR standard deviation by gross tumor size and alternating T variable. **(b)** Scatterplot of NCN ROR calculated using T variable 0 and 1 to illustrate and estimate the effect of the T variable. **(c)** Distributions of NCN ROR by gross tumor size and alternating T variable.

Figure 5a). The isolated effect of tumor size was further illustrated by plotting ROR calculated with T=1 versus ROR calculated with T=0 for the same set of samples and was estimated to 4.61 by linear regression (Supplemental methods Figure 5b). This illustrate how alternating the tumor size variable from 0 to 1, i.e., corresponding to tumor size ≤20 mm or >20mm, results in an increase in ROR by 4.61. The estimated effect of tumor size on ROR is within $2SD_{avg}$ across tumors in the training cohort with gross size >20mm ($SD_{avg}$ 2.16 for T1). This could be expressed as that for the $SD_{avg}$ for T1 tumors the ROR adjustment would be within the 95% range of the ROR distribution from the n=100 different static reference sets. The estimated offset was similar for both subgroups of gross tumor size, 4.60 for T0 and 4.64 for T1. The effect of the tumor size variable on ROR was also illustrated by distributions for respective subgroups of gross tumor size using alternate settings for parameter T (0 or 1) (Supplemental methods Figure 5c). One practical implication of the relatively small direct effect of tumor size is that tumor size would not cause samples to shift from *low* to *high* ROR risk category. Instead, only samples close to the cutoffs are at risk of shifting in or out of the *intermediate* category, e.g., from *low* to *intermediate* or vice versa. We therefore used ROR calculated with T=0 for all samples and transformed the numerical value into categorical using a bin width of 5, i.e., ≤5, 6-10, 11-15, .., 91-95, 96-100) to obtain ROR class-labels for training a SSP model. A minimum of 100 cases was required in a bin to include it in training, resulting in a total of 5359 GEX used in training with the following binned ROR labels: <5 n=100, 6-10 n=186, 11-15 n=272, 16-20 n=314, 21-25 n=324, 26-30 n=317, 31-35 n=361, 36-40 n=318, 41-45 n=311, 46-50 n=364, 51-55 n=384, 56-60 n=390, 61-65 n=411, 66-70 n=366, 71-75 n=349, 76-80 n=272, 81-85 n=196, 86-90 n=124). Accordingly, we trained one SSP-ROR model to use for all tumors irrespectively of size ≤20 mm or >20 mm followed by a simple adjustment for gross tumor size variable. Basically, for cases with tumor size ≤20 mm the SSP-ROR assigned score was used as is, whereas for cases with tumor size >20 mm the SSP-ROR assigned score was adjusted with +5 to appropriately account for the effect of the gross tumor size variable.

*Training SSP models for conventional clinical markers.*

We trained SSP models for the conventional clinical markers: ER, PR, HER2, NHG, and Ki67. Cases in the training cohort with available clinicopathological information were used. Receptor status as determined by respective local pathology department was used. In Sweden, clinical guidelines dictate that tumors with ≤10% of cells with IHC staining for ER and PR status are categorized as negative (Negative). For HER2 status, a case is categorized as negative if IHC staining score <2 or, for patients with IHC 2+ if the ISH status is non-amplified. For training the SSP-ER model, a total of 4786 GEX were used (Negative n=740, and Positive n=4046). The maximum overall agreement was observed at 19 gene-rules per group by AIMS, although only marginal improvements was seen using >3 rules. The union of unique Entrez ID in selected rules was 38. For training the SSP-PR model, a total of 4782 GEX were used (Negative n=1489, and Positive n=3293). The maximum overall agreement was observed already with a single gene-rule that included PR and slowly decreased with increased rules. To retain rule redundancy, we selected to use 3 gene rules. The union of unique Entrez ID in selected rules was 6. For training the SSP-HER2, a total of 4640 GEX were used (Negative n=3986, and Positive n=654). The maximum overall agreement was observed at 8 gene-rules per group. The union of unique Entrez ID in selected rules was 16. For training the SSP-HER2 specific for ER+ cases, a total of 3708 GEX were used (Negative n=3291, and Positive n=417). The maximum overall agreement was observed at 8 gene-rules per group. The union of unique Entrez ID in selected rules was 16. For training the SSP-HER2 specific for ER- cases, a total of 658 GEX were used (Negative n=460, and Positive n=198). The maximum overall agreement was observed at 7 gene-rules per group. The union of unique Entrez ID in selected rules was 14. For training the SSP-Ki67, a total of 3975 GEX were used (High n=2193, and Low n=1782). The maximum overall agreement was observed at 12 gene-rules per group. The union of unique Entrez ID in selected rules was 24. For training the SSP-NHG, a total of 4410 GEX were used (Grade 1 n= 670, Grade 2 n=2191 and Grade 3 n=1549). The maximum overall agreement was observed at 3 gene-rules per group. The union of unique Entrez ID in selected rules was 15.

#### *Time to event analysis*

Primary endpoint for time-to-event analysis was distant recurrence-free interval (DRFi). Additional endpoints were overall survival (OS), recurrence-free interval (RFi) and breast cancer-free interval (BCFi). Distant recurrence free interval was defined as the time from breast cancer diagnosis until distant recurrence. Death was regarded as a censoring event. Overall survival was defined as the time from breast cancer diagnosis until death from any cause. For OS, complete follow-up from the Swedish population registry via NKBC was available until November 16, 2021. Recurrence free interval was defined as the time from breast cancer diagnosis until any recurrence (local, regional or distant). Death was regarded as a censoring event.

For BCFi included events were: i) any breast cancer recurrence (local, regional or distant), ii) a contralateral breast cancer (invasive or in-situ), and iii) breast cancer related death. For a contralateral breast cancer to be regarded as an event we required that it occurred >90 days from diagnosis. If a case was recorded ≤90 days from diagnosis it was not regarded as a separate event and if it was also classified as invasive the case was excluded from survival analysis. Death other than from breast cancer related death was regarded as a censoring event.

In accordance with the standard operating procedures for reporting to the Swedish National Quality Register (NKBC), recurrence events are reported continuously whereas follow-up for disease-free status is only reported at set time-points after diagnosis (at 1 year and at 5 years after diagnosis). To avoid considerable censoring at 1 year with subsequent overestimation of the recurrence rate, the BCFi follow-up time for recurrence-free patients was extended to the follow-up time for overall-survival. However, to minimize false negative rates for recurrence, a time margin to allow for reporting recurrences to NKBC was applied according to the following:

i) For cases with available extended survival time that were event-free and were the extended survival time was >6 months longer than other available NKBC reported follow-up times we reduce the extended follow-up from survival with 6 months (183 days) to have a safety margin.

ii) For cases that were event-free and were we had extended survival time, but the extended survival time was ≤6 months than available NKBC reported follow-up times, we used the available NKBC reported follow-up time. We also required a minimum of 365 days of available follow-up time for overall survival to assure time for healthcare staff to report events to NKBC.

Extending the follow-up for BCFi as outlined above was assessed as feasible by observed recurrence rates that were in line with national Swedish estimates, and by comparison to a large subset of patients where extended journal review was specifically performed to curate recurrence status. No significant difference in recurrence rate was observed for the journal review group.

#### *Emulated treatment recommendation (ETR) in patients with ER+/HER2-/N0 and pT1-2 tumors*

The clinical value of any prognostic stratification is ultimately its influence on disease management such as treatment recommendations. In order to better assess how molecular stratification by, e.g., developed SSP models may affect adjuvant chemotherapy recommendations in a clinical setting we employed a stratification that emulate treatment recommendation (ETR) in patients with ER+/HER2-/N0 and pT1-2 tumors. The ETR schema we used was adapted from available Norwegian guidelines for adjuvant systemic treatment in patients with ER+/HER2-/N0 status when the Prosigna test is available (IS-2945 table 7.4.1 from 08/2020). Our ETR adheres to the actual general recommendations of the Norwegian guidelines but is simplified in that it disregards the guidelines additional considerations for escalation or de-escalation. We employed our ETR to stratify patients into three groups with respect to recommended adjuvant treatment: None, Endo (i.e., endocrine treatment alone for 5-10 years), or ChemoEndo (i.e., adjuvant chemotherapy followed by endocrine treatment for 5-10 years).

The ETR stratification was done as follows:

Tumor size variable pT was assigned using pathology determined tumor size (mm) from NKBC data according to:

Tis = in situ only
T1mi = size ≤1
T1a = size >1 ≤5
T1b = size >5 ≤10
T1c = size >10 ≤20
T2 = size >20 ≤50
T3 = size >50

ER status was categorized using pathology determined ER% staining from NKBC data according to:

ER<50 = ER% staining <50%
ER≥50 = ER% staining ≥50%

Using RNAseq classification for Subtype (LumA, LumB, Her2 or Basal) and ROR risk category (Low, Intermediate, and High) in combination with the pT variable, categorized ER status and pathology determined NHG status from NKBC data we assigned ETR (None, Endo or ChemoEndo) according to:

None = LumA & Low & (T1a OR T1b)
None = LumA & Low & T1c & Grade1
Endo = LumA & Low & T1c & (Grade2 OR Grade3)

Endo = LumA & Low & T2
Endo = LumA & Intermediate & (T1a OR T1b)
Endo = LumA & Intermediate & T1c

Endo = LumA & Intermediate & T2

Endo = (LumB OR Basal OR Her2) & Intermediate & (T1a OR T1b) & ER≥50
ChemoEndo = (LumB OR Basal OR Her2) & Intermediate & (T1a OR T1b) & ER<50

Endo = (LumB OR Basal OR Her2) & Intermediate & T1c & ER≥50
ChemoEndo = (LumB OR Basal OR Her2) & Intermediate & T1c & ER<50

ChemoEndo = (LumB OR Basal OR Her2) & Intermediate & T2 & ER≥50
ChemoEndo = (LumB OR Basal OR Her2) & Intermediate & T2 & ER<50

Endo = High & (T1a OR T1b) & ER≥50
ChemoEndo = High & (T1a OR T1b) & ER<50

ChemoEndo = High & T1c
ChemoEndo = High & T2

All others unassigned.

#### *Statistics*

All p-values reported from statistical tests are two-sided if not otherwise specified. Box-plot elements corresponds to: i) center line = median, ii) box limits = upper and lower quartiles, iii) whiskers = 1.5x interquartile range.

### *References*

1. Vallon-Christersson, J., Nordborg, N., Svensson, M. & Hakkinen, J. BASE--2nd generation software for microarray data management and analysis. *BMC Bioinformatics* **10**, 330 (2009).
2. UCSC Genome Browser. http://genome.ucsc.edu/. Accessed:
3. dbSNP. https://www.ncbi.nlm.nih.gov/SNP/. Accessed: 2021. Nov 12
4. Frankish, A., *et al.* GENCODE reference annotation for the human and mouse genomes. *Nucleic acids research* **47**, D766-D773 (2019).
5. Picard tools. https://broadinstitute.github.io/picard/. Accessed: 2021. Nov 12
6. Bolger, A.M., Lohse, M. & Usadel, B. Trimmomatic: a flexible trimmer for Illumina sequence data. *Bioinformatics* **30**, 2114-2120 (2014).
7. Langmead, B. & Salzberg, S.L. Fast gapped-read alignment with Bowtie 2. *Nature methods* **9**, 357-359 (2012).
8. Kim, D., Paggi, J.M., Park, C., Bennett, C. & Salzberg, S.L. Graph-based genome alignment and genotyping with HISAT2 and HISAT-genotype. *Nature biotechnology* **37**, 907-915 (2019).
9. Pertea, M., *et al.* StringTie enables improved reconstruction of a transcriptome from RNA-seq reads. *Nature biotechnology* **33**, 290-295 (2015).
10. Parker, J.S., *et al.* Supervised risk predictor of breast cancer based on intrinsic subtypes. *J Clin Oncol* **27**, 1160-1167 (2009).
11. Lauss, M., *et al.* Monitoring of technical variation in quantitative high-throughput datasets. *Cancer Inform* **12**, 193-201 (2013).
12. Wilkerson, M.D. & Hayes, D.N. ConsensusClusterPlus: a class discovery tool with confidence assessments and item tracking. *Bioinformatics* **26**, 1572-1573 (2010).
13. Gnant, M., *et al.* Predicting distant recurrence in receptor-positive breast cancer patients with limited clinicopathological risk: using the PAM50 Risk of Recurrence score in 1478 postmenopausal patients of the ABCSG-8 trial treated with adjuvant endocrine therapy alone. *Ann Oncol* **25**, 339-345 (2014).
14. Filipits, M., *et al.* The PAM50 risk-of-recurrence score predicts risk for late distant recurrence after endocrine therapy in postmenopausal women with endocrine-responsive early breast cancer. *Clin Cancer Res* **20**, 1298-1305 (2014).
15. Wallden, B., *et al.* Development and verification of the PAM50-based Prosigna breast cancer gene signature assay. *BMC Med Genomics* **8**, 54 (2015).
16. Paquet, E.R. & Hallett, M.T. Absolute assignment of breast cancer intrinsic molecular subtype. *Journal of the National Cancer Institute* **107**, 357 (2015).
17. Saal, L.H., *et al.* The Sweden Cancerome Analysis Network - Breast (SCAN-B) Initiative: a large-scale multicenter infrastructure towards implementation of breast cancer genomic analyses in the clinical routine. *Genome Med* **7**, 20 (2015).

**Supplement methods Table 1.** GEX cohorts balanced for clinical subgroups for constructing the library protocol adjustment procedure.

**COHORT 1**
n=1988
dUTP/TruSeq

| Clinical subgroup | dUTP | TruSeq |
|---|---|---|
| ERpHER2nLNn_Cyto | 2 | 2 |
| ERpHER2nLNn_Endo | 346 | 346 |
| ERpHER2nLNn_EndoCyto | 100 | 100 |
| ERpHER2nLNn_ImmuEndoCyto | 2 | 2 |
| ERpHER2nLNn_NA | 1 | 1 |
| ERpHER2nLNn_None | 69 | 69 |
| ERpHER2nLNp_Endo | 130 | 130 |
| ERpHER2nLNp_EndoCyto | 132 | 132 |
| ERpHER2nLNp_ImmuEndoCyto | 2 | 2 |
| ERpHER2nLNp_None | 5 | 5 |
| HER2pERn_ImmuCyto | 31 | 31 |
| HER2pERn_ImmuEndoCyto | 1 | 1 |
| HER2pERn_NA | 2 | 2 |
| HER2pERn_None | 9 | 9 |
| HER2pERp_Endo | 11 | 11 |
| HER2pERp_EndoCyto | 4 | 4 |
| HER2pERp_ImmuCyto | 4 | 4 |
| HER2pERp_ImmuEndoCyto | 56 | 56 |
| HER2pERp_NA | 1 | 1 |
| HER2pERp_None | 3 | 3 |
| NA_Cyto | 1 | 1 |
| NA_Endo | 1 | 1 |
| TNBC_Cyto | 57 | 57 |
| TNBC_Endo | 2 | 2 |
| TNBC_EndoCyto | 1 | 1 |
| TNBC_NA | 2 | 2 |
| TNBC_None | 19 | 19 |

**COHORT 2**
n=1824
dUTP/NeoPrep

| Clinical subgroup | dUTP | NeoPrep |
|---|---|---|
| ERpHER2nLNn_Cyto | 1 | 1 |
| ERpHER2nLNn_Endo | 331 | 331 |
| ERpHER2nLNn_EndoCyto | 100 | 100 |
| ERpHER2nLNn_ImmuEndoCyto | 1 | 1 |
| ERpHER2nLNn_NA | 2 | 2 |
| ERpHER2nLNn_None | 67 | 67 |
| ERpHER2nLNp_Endo | 102 | 102 |
| ERpHER2nLNp_EndoCyto | 132 | 132 |
| ERpHER2nLNp_None | 1 | 1 |
| HER2pERn_ImmuCyto | 21 | 21 |
| HER2pERn_NA | 1 | 1 |
| HER2pERn_None | 3 | 3 |
| HER2pERp_Endo | 9 | 9 |
| HER2pERp_EndoCyto | 1 | 1 |
| HER2pERp_ImmuEndoCyto | 52 | 52 |
| HER2pERp_NA | 1 | 1 |
| NA_Cyto | 1 | 1 |
| TNBC_Cyto | 57 | 57 |
| TNBC_Endo | 1 | 1 |
| TNBC_EndoCyto | 2 | 2 |
| TNBC_ImmuCyto | 1 | 1 |
| TNBC_NA | 2 | 2 |
| TNBC_None | 23 | 23 |

**COHORT 3**
n=1852
NeoPrep/TruSeq

| Clinical subgroup | NeoPrep | TruSeq |
|---|---|---|
| ERpHER2nLNn_Cyto | 1 | 1 |
| ERpHER2nLNn_Endo | 331 | 331 |
| ERpHER2nLNn_EndoCyto | 102 | 102 |
| ERpHER2nLNn_ImmuEndoCyto | 1 | 1 |
| ERpHER2nLNn_NA | 1 | 1 |
| ERpHER2nLNn_None | 67 | 67 |
| ERpHER2nLNp_Endo | 102 | 102 |
| ERpHER2nLNp_EndoCyto | 143 | 143 |
| ERpHER2nLNp_None | 1 | 1 |
| HER2pERn_ImmuCyto | 21 | 21 |
| HER2pERn_NA | 1 | 1 |
| HER2pERn_None | 3 | 3 |
| HER2pERp_Endo | 9 | 9 |
| HER2pERp_EndoCyto | 1 | 1 |
| HER2pERp_ImmuEndoCyto | 52 | 52 |
| HER2pERp_NA | 1 | 1 |
| NA_Cyto | 1 | 1 |
| TNBC_Cyto | 65 | 65 |
| TNBC_Endo | 1 | 1 |
| TNBC_EndoCyto | 1 | 1 |
| TNBC_NA | 2 | 2 |
| TNBC_None | 19 | 19 |
